# The Effect of Firearm Laws on Pediatric Mortality in the United States

**DOI:** 10.1101/2024.09.21.24314130

**Authors:** Jeremy Samuel Faust, Ji Chen, Shriya Bhat, Onyekachi Otugo, Benjamin Renton, Alexander Junxiang Chen, Zhenqiu Lin, Harlan M. Krumholz

## Abstract

**Introduction:** Firearms are the leading cause of death in US children and adolescents, but little is known about whether legal policies may be responsible.

**Methods:** We conducted difference-in-differences analysis on CDC WONDER data before and after *McDonald v. Chicago*, the landmark 2010 Supreme Court decision on firearms regulation. States were divided into three groups, based on legal actions taken before and since 2010, most permissive, permissive, and restricted. Firearm mortality trends before (1999-2010) and after (2010-2023) were determined and compared across the three groups for all intents and by intent (homicide and suicide). Within the most permissive state grouping, pediatric firearm mortality by 2013 urbanicity and by observed race and ethnicity were conducted. For each US state, pre-and- post 2010 all-intent pediatric firearm mortality incident rates were compared.

**Results:** There were 7130 excess pediatric firearms deaths in states with more permissive regulatory regimes than those with stricter frameworks, as well as higher rates of homicide and suicide. Non-Hispanic Black populations were disproportionately affected by these trends. Four states (California, Maryland, New York, and Rhode Island) had decreased pediatric firearm mortality after 2010, all of which were in the restrictive firearms law group.

**Conclusion:** States with more permissive firearm laws have experienced greater pediatric firearm mortality during the post-*McDonald v. Chicago* era.

**KEY POINTS:** 

**Question:** Did states enacting permissive firearm laws after 2010—when *McDonald v. Chicago* was decided by the United States Supreme Court—subsequently experience higher rates of pediatric firearm mortality?

**Findings:** Difference-in-difference analysis found that state groups that enacted more permissive firearm laws after 2010 experienced >7,100 firearm deaths in children and adolescents ages 0-17 between 2010-2023 compared to restrictive law-enacting states, of which most (77.5%) were homicides. In the permissive states groups, increases occurred in all urbanicities. The largest increase occurred in non-Hispanic Black children and adolescents. Four states had statistical decreases in pediatric firearm mortality during the study period, all of which were in states which enacted restrictive firearm policies.

**Meaning:** Permissive firearm laws contributed thousands of excess firearm deaths among children living in states with permissive policies. Future work should focus on determining which types of laws conferred the most harm and which the most protection.

## Introduction

Firearm deaths are now the leading cause of death among US children and adolescents.^1^ Permissive firearm laws are associated with higher all-age firearm-related mortality.^2^ (tk). In 2010, the US Supreme Court decided *McDonald v. Chicago*, which applied the Second Amendment to the states.^3^ After *McDonald*, many US states subsequently changed their firearm purchase and other firearm-related use laws and restrictions. This study investigates whether increases in pediatric firearm mortality were linked to more permissive firearm laws.

## Methods

### State groupings

We categorized US states into three groups: most permissive, permissive, and strict based on legal changes since 2010 (Table S1-S2). The classification of states firearm ownership and use policies was adapted from a composite score synthesizing three established metrics (for details see Supplemental Methods).

### Difference in Difference analysis

We estimated the association between the effects of *McDonald v Chicago* by conducting a difference-in-differences analysis of pre- (1999-2010) and post-policy (2010-2023) mortality slopes in comparison to other state groupings.^4^ For state grouping rationales, see Table S1-2.

Analyses were conducted for all-intent firearm mortality and by intent, respectively. Specifically, we assume that the mortality rate *p*_*ij*_ for state group *i* at time *j* follows a logistic regression, and the model incorporates time (yearly), state group (by legal status; see Table S1-2), a time-varying variable indicating the time since the policy enacted (2010), and the interaction term of each state group and the time-varying variable; that is,

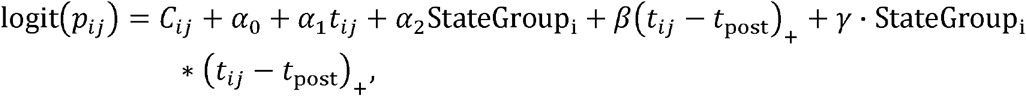

and *t*_+_ = max {*t*, 0}, *C*_*i*_ is a known offset term for State group *i*. In the model, (*t*_*ij*_ - *t*_*post*_)_+_ is to evaluate the change in mortality rate slope after the *McDonald* decision; the interaction term StateGroup_i_ * (*t*_*ij*_ - *t*_*post*_)_+_ is to evaluate whether the post-decision change in mortality rate slope for each state group differs from the reference group (Strict states group). The difference is considered significant if the interaction p-value < 0.05.

Subgroup analyses by 2013 urbanicity and by race and ethnicity for groups without suppression (non-Hispanic White, non-Hispanic Black or African American, and Hispanic) were conducted separately within a state group, using the same model and adjusting for the respective subgroup variable. Race and ethnicity were determined from CDC WONDER (Wide-ranging ONline Data for Epidemiologic Research), which reports observed race and ethnicity at the time of death.^5^

After model fitting, the observed mortality rate and counterfactual mortality rate (assuming the trend in each state group would be the same as that in the strict state group reference group during the post-*McDonald* period) are estimated for each year for all firearm deaths, and by intent; for the urbanicity and race/ethnicity analysis (which assessed only the most permissive firearm laws group of states), the counterfactual was projected expected deaths, assuming the trend in each group would not change after *McDonald*. The excess mortality rate for each year is then calculated as the observed rate minus the counterfactual rate. Subsequently, for each state group, the total number of excess deaths and the overall excess mortality rate in the post- *McDonald* period are calculated, taking into account the population.

### State analysis

The pre- and post-*McDonald* pediatric firearm incident rates (all intents) were measured for each state (except Hawaii, which had inadequate data due to small numbers). For each state, an incident rate ratio comparing the pediatric firearm mortality before (1999-2010) and after (2010-2023) was measured, with their corresponding 95% confidence intervals. IRRs were considered significant if they did not include 1.0.

This study followed the Strengthening the Reporting of Observational Studies in Epidemiology (STROBE) guidelines and was exempt from institutional review board consideration because it used public data.

## Results

During the overall study (1999-2023), there were an average of 73.3 million US residents ages 0- 17 (1.8 billion person-years) in whom death certificate data were available in the 1,015,491 deaths described in CDC WONDER; mean decedent age [SD] for 3.9 years [6.1] for all-cause pediatric mortality and 14.4 years [3.8] for pediatric firearm-specific mortality (all intents). There were 41,269 pediatric firearm deaths during the study period, accounting for 4.1% of all pediatric deaths.

Compared with states with strict firearm laws, from 2010-2023 there were a combined 7,130 excess deaths due to firearms in the most permissive and permissive states; 5,893 excess deaths (1.19 per 100,000 children, interaction p<0.001) in the most permissive group, and 1,236 excess deaths (0.72 per 100,000 children, interaction p<0.001) in the permissive group (Figure, Panel A, Figure S1). Sensitivity analyses showed decreased mortality rates or insignificant differences in other causes of death (Figure S2-3).

Pediatric firearm mortality increased for homicide (∼77.5% of excess firearm deaths) and suicide in permissive states (Figure, Panel B-C). In the most permissive states, mortality occurred across all urbanicities (Panel D) and was highest among non-Hispanic Black or African American populations (Panel E).

Compared to the pre-*McDonald* period, pediatric firearm mortality (all intents) increased in 33 of 49 states (67.3%) with adequate data. New Hampshire (IRR 2.0; 95% CI 1.1-3.6) and New Hampshire (IRR 1.8; 95% CI 1.5-2.1) had the largest relative increases, both of which were in the most permissive state firearm laws state grouping. Four states had statistically significant decreases during the study period, all of which were in the restrictive firearm laws state grouping. Rhode Island (IRR 0.4; 95% CI 0.2-0.8) and California (IRR 0.6; 95% CI 0.6-0.7) had the largest relative decreases.

## Discussion

This report suggests that more permissive gun ownership laws were responsible for increases in firearm mortality among US children. Increases occurred in both homicide and suicide, but primarily by homicide, outpacing the usual predominance of homicide as the leading intent in firearm deaths among the non-adult population.^6,7^

In the most permissive firearm laws state grouping, increases in pediatric firearm mortality occurred in all urbanicity categories, despite reports that burdens in overall (all ages) firearm mortality may be increasing in rural areas.^8^

Increased firearm deaths among non-Hispanic Black populations may reflect disproportionate increases in firearm ownership during the study.^9,10^ Inconsistent physician adherence (by patient race/ethnicity) and the effectiveness of received anticipatory guidance—related to safe storage, for example—may be an explanation, as observed previously regarding car safety recommendations.^11^

States have enacted several types of firearm laws, though it remains unknown which of these laws may be associated or directly responsible for increases or decreases in pediatric firearm mortality. For example, safe storage laws are aimed at decreasing accidental firearm deaths, especially for the >30 million US children who live in homes with a firearm. However, accidental deaths are a small proportion of pediatric firearm mortality, implying that other policies may have larger impact.^12,13^ Some laws such as stand-your-ground have been associated with all-ages increases in firearm mortality, but not specifically in the non-adult population.^14^ Other laws, including universal background checks, have been associated with all ages reductions in firearm mortality.^15^ Notably, however, pediatric firearm deaths decreased in several states that enacted strict firearm laws during the post-*McDonald* period. Therefore, future work should determine which types of laws (e.g., safe storage, required safety training for gun owners) and which evidence-based practices reduce pediatric firearm deaths, potentially informing evidence-based policies to reduce pediatric firearm mortality, with the goal of bringing the United States into line with lower pediatric (and all ages) firearm mortality with those of peer nations.^16–18^

This study has limitations. First, we used the 2010 *McDonald* decision as the barrier for the difference-in-difference analysis, rather than the specific dates when firearm laws in the states were enacted. This was chosen for data clarity. Second, while difference-in-difference inherently controls for other factors that would not be expected to change across the pre-and-post periods, certainty is not possible. However, the groupings were such that the findings are not likely to be ecologic, as similar changes in other causes of death assessed in the sensitivity analyses were not observed, and state groupings did not merely replicate geographic regions. Third, there was not enough data to assess smaller race and ethnicity groupings.

In conclusion, this study demonstrates that substantial increases in pediatric firearm mortality in the last 15 years are linked to state-level legal policies. States with permissive firearm laws experienced thousands more pediatric deaths than would have occurred had their post-*McDonald* firearm mortality trends matched those in states with restrictive firearm laws, indicating that these deaths are not inevitable.

## Data Availability

All data analyzed in this study are derived from the CDC WONDER database.

---

**Figure.**
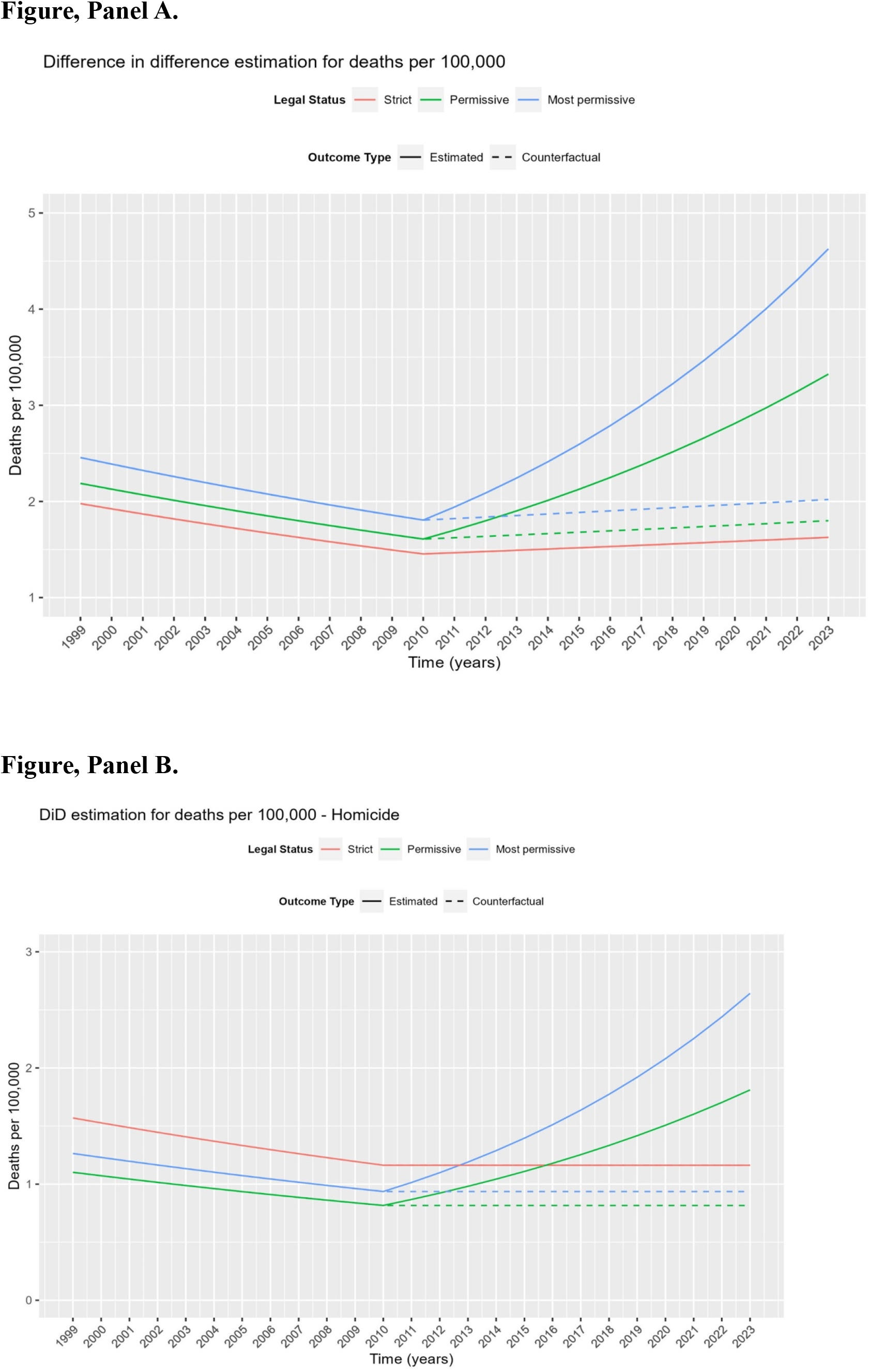
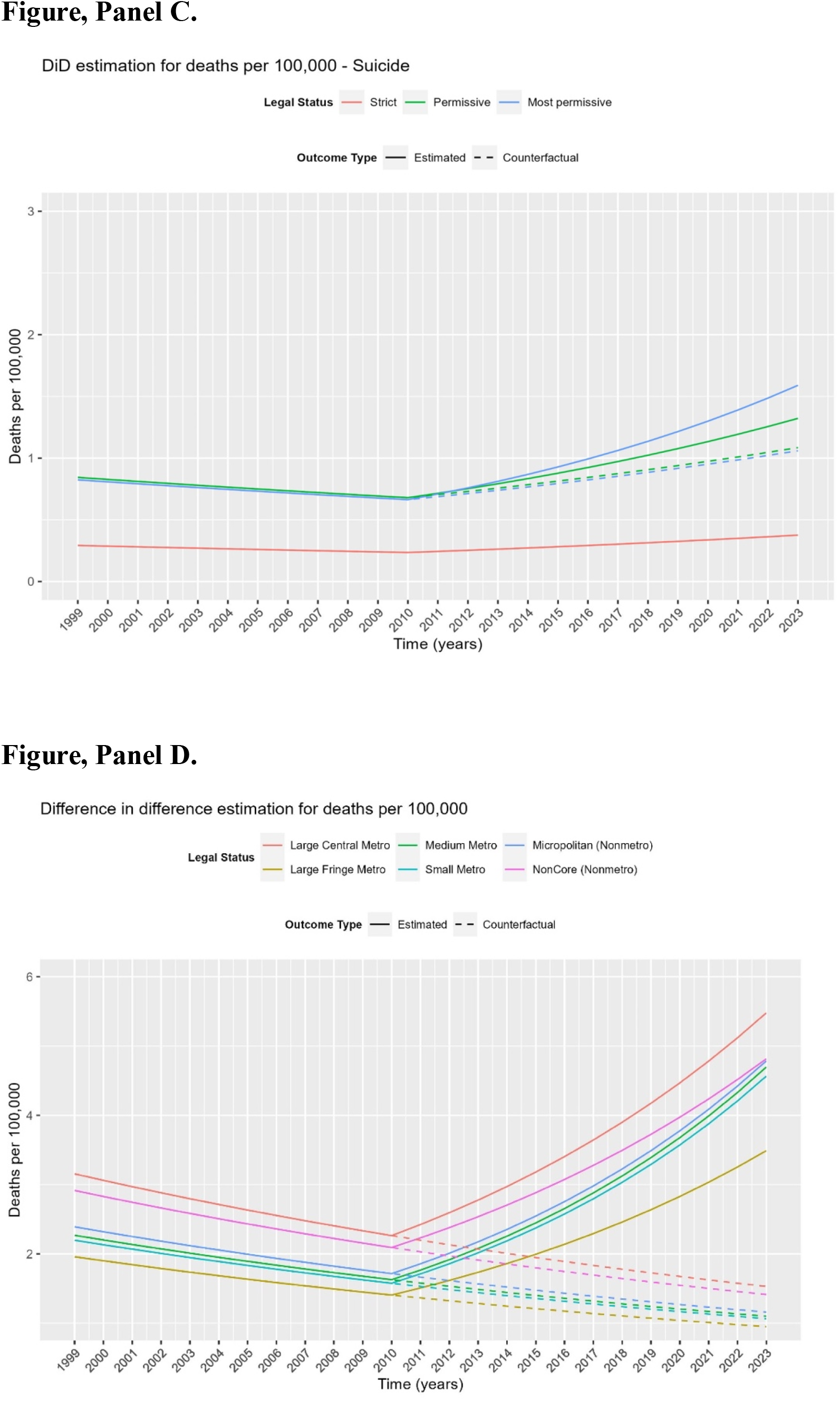
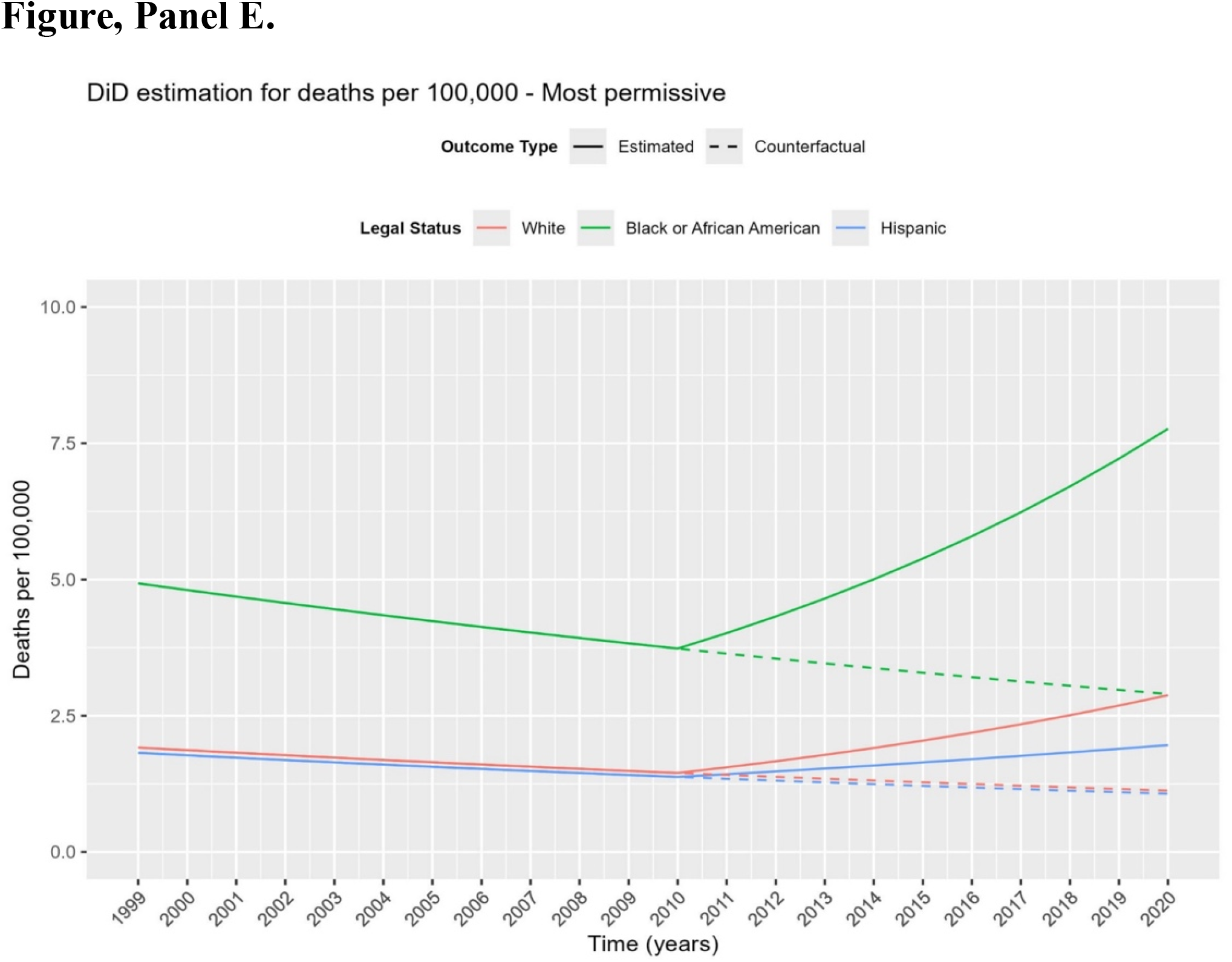
Pediatric firearm deaths before and after *McDonald v. Chicago* (2010). Solid lines indicate mortality slopes as determined by logistic regression of yearly data before and after 2010. Dashed lines indicate expected deaths, using the post-2010 slope in the strict states group as the counterfactual for the permissive and most permissive states group. Panel A: Pediatric firearm deaths, all intents, by legal grouping. Blue lines indicate the group of states with most permissive firearm laws after *McDonald*, green lines indicate the group of states with moderately permissive firearm laws, and red lines indicate the group of states with strict firearm laws, 1999-2023. Panel B: Pediatric firearm deaths, homicide, by legal grouping, 1999-2023. Panel C: Pediatric firearm deaths, suicide, by legal grouping, 1999-2023. Panel D: Pediatric firearm deaths, all intents, by 2013 urbanicity in the most permissive states group, 1999-2023. Panel E: Pediatric firearm deaths, all intents, by race/ethnicity in the most permissive states group, 1999-2020. Note: 2021-2023 was not assessed due to changes in CDC reporting categories.

**Table.** Incident rates pre- and post-*McDonald*, and incident rate ratios. Red indicates statistically significant increase. Blue indicates statistically significant decrease.

| State | Legal grouping | Firearm mortality, crude rate 1999-2010 | Firearm mortality, crude rate 2010-2023 | Incident rate ratio (95% confidence interval) |
| --- | --- | --- | --- | --- |
| Alabama | Most permissive | 2.8 | 3.9 | 1.4 (1.2-1.6) |
| Alaska | Most permissive | 6.1 | 5.4 | 0.9 (0.7-1.1) |
| Arizona | Most permissive | 2.7 | 2.4 | 0.9 (0.8-1.0) |
| Arkansas | Most permissive | 2.8 | 3.9 | 1.4 (1.2-1.6) |
| Florida | Most permissive | 1.8 | 2.4 | 1.4 (1.2-1.5) |
| Georgia | Most permissive | 2.1 | 3.2 | 1.5 (1.4-1.7) |
| Idaho | Most permissive | 2.4 | 3.0 | 1.3 (1.0-1.6) |
| Indiana | Most permissive | 2.0 | 2.9 | 1.5 (1.3-1.6) |
| Iowa | Most permissive | 1.4 | 1.8 | 1.4 (1.1-1.7) |
| Kansas | Most permissive | 1.9 | 3.0 | 1.6 (1.3-2.0) |
| Kentucky | Most permissive | 1.7 | 3.1 | 1.8 (1.5-2.1) |
| Louisiana | Most permissive | 4.1 | 5.7 | 1.4 (1.3-1.6) |
| Maine | Most permissive | 0.9 | 1.4 | 1.5 (1.0-2.3) |
| Mississippi | Most permissive | 3.4 | 5.1 | 1.5 (1.3-1.7) |
| Missouri | Most permissive | 2.9 | 4.2 | 1.4 (1.3-1.6) |
| Montana | Most permissive | 3.7 | 4.2 | 1.1 (0.9-1.5) |
| Nebraska | Most permissive | 2.1 | 1.8 | 0.8 (0.6-1.1) |
| New Hampshire | Most permissive | 0.5 | 0.9 | 2.0 (1.1-3.6) |
| North Carolina | Most permissive | 2.1 | 2.8 | 1.4 (1.2-1.5) |
| North Dakota | Most permissive | 2.5 | 2.8 | 1.1 (0.8-1.7) |
| Ohio | Most permissive | 1.6 | 2.8 | 1.7 (1.6-1.9) |
| Oklahoma | Most permissive | 2.6 | 3.6 | 1.4 (1.2-1.6) |
| South Carolina | Most permissive | 2.3 | 3.9 | 1.7 (1.5-1.9) |
| South Dakota | Most permissive | 2.8 | 2.4 | 0.9 (0.6-1.2) |
| Tennessee | Most permissive | 2.4 | 3.6 | 1.5 (1.3-1.7) |
| Texas | Most permissive | 1.9 | 2.5 | 1.3 (1.2-1.4) |
| Utah | Most permissive | 1.3 | 2.3 | 1.7 (1.4-2.1) |
| West Virginia | Most permissive | 2.0 | 2.2 | 1.1 (0.9-1.5) |
| Wisconsin | Most permissive | 2.0 | 2.3 | 1.1 (1.0-1.3) |
| Wyoming | Most permissive | 3.0 | 4.2 | 1.4 (1.0-2.0) |
| Colorado | Permissive | 1.8 | 3.0 | 1.7 (1.4-1.9) |
| Delaware | Permissive | 1.5 | 1.9 | 1.3 (0.8-1.9) |
| Michigan | Permissive | 2.2 | 2.5 | 1.1 (1.0-1.3) |
| Minnesota | Permissive | 1.4 | 1.6 | 1.1 (1.0-1.4) |
| Nevada | Permissive | 2.3 | 2.7 | 1.2 (1.0-1.4) |
| New Mexico | Permissive | 3.4 | 3.9 | 1.2 (1.0-1.4) |
| Oregon | Permissive | 1.3 | 1.8 | 1.4 (1.1-1.7) |
| Pennsylvania | Permissive | 1.9 | 2.5 | 1.3 (1.2-1.5) |
| Vermont | Permissive | 1.1 | 1.8 | 1.7 (0.9-3.0) |
| Virginia | Permissive | 2.0 | 2.4 | 1.2 (1.1-1.3) |
| Washington | Permissive | 1.5 | 1.8 | 1.2 (1.1-1.4) |
| California | Strict | 2.2 | 1.4 | 0.6 (0.6-0.7) |
| Connecticut | Strict | 0.8 | 1.1 | 1.5 (1.1-1.9) |
| Illinois | Strict | 2.5 | 3.4 | 1.4 (1.2-1.5) |
| Maryland | Strict | 2.6 | 2.0 | 0.8 (0.7-0.9) |
| Massachusetts | Strict | 0.7 | 0.6 | 0.8 (0.6-1.0) |
| New Jersey | Strict | 0.9 | 0.9 | 1.0 (0.8-1.2) |
| New York | Strict | 1.2 | 0.9 | 0.8 (0.7-0.9) |
| Rhode Island | Strict | 1.2 | 0.5 | 0.4 (0.2-0.8) |

\*Hawaii was not included due to data suppression owing to low rates.

