## Supplemental Information for "The Effect of Firearm Laws on Pediatric Mortality in the United States"

**Supplement Material for Faust et al, “The Effect of Firearm Laws on Pediatric Mortality in the United States.:**

**Supplemental methods**

**Legal classifications.** States were classified into one of three groups (most permissive, permissive, and restrictive) based on existing and post-2010 enacted firearm ownership and use policies using a rubric adapted from a composite score synthesizing three established metrics: Giffords Law Center grades, Everytown for Gun Safety scores, and Brady Campaign scores. These individual measures were normalized to a uniform 0-1 scale to ensure comparability using the min-max scaling method, also known as feature scaling. A composite score was then computed for each state by averaging these normalized values. States were subsequently classified into three categories based on their composite scores: "restrictive" (scores ≥ 0.66), "less permissive" (0.33 ≤ scores < 0.66), and "permissive" (scores < 0.33). This tripartite classification scheme aims to provide a comprehensive representation of firearm policy stringency across states, condensing multiple commonly used assessments into a single, easily interpretable metric."

**More on the normalization:** 
The normalization process in this analysis was conducted using the min-max scaling method, also known as feature scaling. For each of the three metrics (Giffords Numeric, Everytown for Gun Safety Score, and Brady Campaign Scores), the following formula was utilized:

$$\eta= (x - min(\chi)) / (max(\chi) - min(\chi))$$

Where x is the original value, min(x) is the minimum value in the dataset for that metric, and max(x) is the maximum value in the dataset for that metric. This transformation is applied to each value within each metric column, where the lowest value becomes 0, the highest value becomes 1, and all other values are scaled proportionally between 0 and 1. The composite score for each state was calculated by taking the average of the three normalized firearm ownership and use metrics.

**Supplement exhibits**

**Table S1**

| Alabama | 2006 (L): Stand Your Ground Law - Removed duty to retreat before using deadly force in self-defense situations. 2013 (L): Workplace Protection Act - Allowed employees to keep firearms in their vehicles at work, overriding employer bans. 2014 (L): Open Carry Clarification - Clarified and expanded open carry rights in the state. 2015 (L): Pistol Permit Reform - Streamlined the pistol permit process and made permits valid for five years. 2019 (L): Church Carry - Allowed churches to establish their own armed security programs. 2022 (L): Constitutional Carry - Implemented permitless carry, allowing legal gun owners to carry concealed firearms without a permit. 2023 (L): Second Amendment Protection Act - Prohibited state and local law enforcement from enforcing certain federal gun laws. 2024 (L): School Guardian Program - Allowed trained school employees to carry firearms on school grounds (Note: If not enacted, this was a significant proposal during this period). |
| --- | --- |
| Alaska | 2003 (L): Constitutional Carry - Alaska became the first state to allow permitless concealed carry for residents 21 and older. 2006 (L): Stand Your Ground Law - Removed the duty to retreat before using deadly force in self-defense situations. 2010 (L): Firearms Freedom Act - Declared that firearms made and kept in Alaska are not subject to federal regulations. 2013 (L): HB 69 - Nullification attempt of federal gun laws, declaring certain federal firearm laws unenforceable in Alaska. 2016 (L): SB 1441 - Prohibited state and municipal entities from using resources to implement federal gun regulations. 2021 (L): HB 186 - Designated firearms and ammunition businesses as "essential" during states of emergency. |
| Arizona | 2006 (L): Castle Doctrine - Expanded self-defense rights in one's home and vehicle. 2009 (L): Firearms in Restaurants - Allowed concealed carry permit holders to bring firearms into restaurants serving alcohol. 2010 (L): Constitutional Carry - Implemented permitless carry, allowing legal gun owners to carry concealed firearms without a permit. 2013 (L): Second Amendment Protection Act - Prohibited state agencies from enforcing new federal gun control measures. 2016 (L): Preemption Strengthening - Enhanced state preemption of local firearm regulations. 2017 (L): Firearms Transfer Protection - Prohibited requiring background checks on private party gun sales. 2021 (L): Second Amendment Sanctuary - Declared Arizona a Second Amendment sanctuary state, further limiting enforcement of federal gun laws. 2022 (L): Young Adult Carry - Allowed individuals aged 18-20 to obtain a provisional concealed carry permit. 2023 (L): School Guardian Program - Allowed trained school staff to carry firearms on school grounds (Note: If not enacted, this was a significant proposal during this period). |
| Arkansas | 2013 (L): Act 746 - Redefined "journey" to allow carrying firearms in vehicles without a permit. 2015 (L): Act 1078 - Expanded concealed carry rights to include certain areas of public college campuses. 2017 (L): Enhanced Carry Permit - Created a new enhanced permit allowing carry in more locations, including some government buildings and bars. 2017 (L): Act 562 - Campus Carry Law - Allowed concealed carry permit holders to carry firearms on public college campuses. 2019 (L): Act 472 - Reduced the age requirement for concealed carry permits from 21 to 18 for active duty military and National Guard members. 2021 (L): Act 1024 - "Stand Your Ground" law - Removed the duty to retreat before using deadly force in self-defense situations. 2021 (L): Act 250 - Prohibited state and local officials from enforcing federal gun restrictions enacted after January 1, 2021. |
| California | 2000 (S): Unsafe Handgun Act - Established safety standards for handguns sold in California. 2001 (S): Handgun Safety Certificate Program - Required handgun buyers to obtain a safety certificate. 2007 (S): Microstamping Law - Required new semi-automatic pistols to have microstamping technology. 2014 (S): Gun Violence Restraining Order Law - Allowed family members and law enforcement to petition courts to remove firearms from individuals deemed a risk. 2016 (S): "Bullet Button" Ban - Expanded assault weapon definition to include certain semi-automatic rifles with detachable magazines. 2016 (S): Large-Capacity Magazine Ban - Prohibited possession of magazines capable of holding more than 10 rounds. 2019 (S): Ammunition Purchase Background Checks - Required background checks for ammunition purchases. 2019 (S): Gun Purchase Limit - Restricted handgun purchases to one per 30-day period (extended to long guns in 2021). 2020 (S): Ghost Gun Regulations - Required background checks for "ghost gun" kits and parts. 2022 (S): Firearm Industry Responsibility Act - Allowed lawsuits against gun manufacturers for irresponsible marketing practices. 2023 (S): Concealed Carry Restrictions - Implemented new restrictions on concealed carry permits in response to NYSRPA v. Bruen. |
| Colorado | 2003 (L): Concealed Carry Act - Made Colorado a "shall issue" state for concealed carry permits. 2007 (L): Castle Doctrine - Expanded self-defense rights in one's home. 2012 (L): Constitutional Carry for Travel - Allowed anyone who can legally possess a firearm to carry it in their vehicle for lawful protection. 2013 (S): Universal Background Checks - Required background checks for private gun sales and transfers. 2013 (S): Large-Capacity Magazine Ban - Prohibited the sale, transfer, and possession of magazines capable of holding more than 15 rounds. 2013 (S): Domestic Violence Gun Ban - Required domestic abusers subject to protection orders to relinquish their firearms. 2019 (S): Red Flag Law - Allowed courts to temporarily remove firearms from individuals deemed a risk to themselves or others. 2021 (S): Safe Storage Law - Required gun owners to securely store firearms when not in use. 2021 (S): Lost or Stolen Firearm Reporting - Required gun owners to report lost or stolen firearms within five days. 2023 (S): Waiting Period - Implemented a three-day waiting period for firearm purchases. 2023 (S): Expanded Red Flag Law - Allowed more parties to petition for extreme risk protection orders. |
| Connecticut | 2013 (S): Gun Violence Prevention and Children's Safety Act - Comprehensive gun control package passed in response to the Sandy Hook shooting, including: Expanded assault weapons ban Ban on magazines holding more than 10 rounds Universal background checks Gun offender registry Expanded requirements for obtaining a firearms permit 2016 (S): Domestic Violence Gun Law - Required individuals subject to temporary restraining orders to surrender firearms within 24 hours. 2018 (S): Bump Stock Ban - Prohibited the sale, purchase, and possession of bump stocks and other rapid-fire devices. 2019 (S): Ghost Gun Ban - Prohibited the manufacture, sale, and possession of ghost guns (unserialized and untraceable firearms). 2019 (S): Safe Storage Law - Strengthened requirements for safe storage of firearms in homes with minors. 2021 (S): Risk Warrant Expansion - Expanded the state's risk warrant (red flag) law to allow family members and medical professionals to petition for temporary firearm removal. 2022 (S): Ghost Gun Law Enhancement - Strengthened existing ghost gun regulations and increased penalties for violations. 2023 (S): Assault Weapon Definition Expansion - Broadened the definition of assault weapons to include more firearm models. |
| Delaware | 2013 (S): Lost or Stolen Firearm Reporting - Required gun owners to report lost or stolen firearms within 7 days. 2015 (S): Gun Relinquishment in Protective Orders - Required individuals subject to protective orders to surrender firearms. 2018 (S): Extreme Risk Protection Order (Red Flag Law) - Allowed courts to temporarily remove firearms from individuals deemed a risk. 2018 (S): Bump Stock Ban - Prohibited the sale, purchase, and possession of bump stocks and other rapid-fire devices. 2018 (S): Age Restriction - Raised the minimum age to purchase or own a firearm from 18 to 21, with some exceptions. 2019 (S): Background Checks for Private Sales - Required background checks for private gun sales and transfers. 2021 (S): Large-Capacity Magazine Ban - Prohibited magazines capable of holding more than 17 rounds. 2022 (S): Assault Weapons Ban - Prohibited the manufacture, sale, and possession of certain semi-automatic firearms defined as assault weapons. 2022 (S): Handgun Qualification License - Required individuals to obtain a permit before purchasing a handgun. 2023 (S): Ghost Gun Ban - Prohibited the manufacture, sale, and possession of unserialized firearms and unfinished frames or receivers. |
| Florida | 2005 (L): Stand Your Ground Law - Removed the duty to retreat before using force in self-defense situations. 2008 (L): Guns at Work Law - Allowed employees to keep firearms in their vehicles at work. 2011 (L): Firearm Preemption Enhancement - Strengthened state preemption of firearms laws, imposing penalties on local officials who enact stricter gun regulations. 2014 (L): Warning Shot Law - Extended Stand Your Ground protections to include the firing of warning shots. 2018 (S): Marjory Stoneman Douglas High School Public Safety Act - Implemented several gun control measures in response to the Parkland shooting: Raised minimum age to purchase firearms from 18 to 21 Instituted a 3-day waiting period for firearm purchases Banned bump stocks Established red flag law (Risk Protection Orders) 2019 (L): Classroom Carry - Allowed teachers to carry firearms in classrooms after completing training. 2021 (L): Church Carry - Allowed concealed carry permit holders to carry firearms in places of worship. 2022 (L): Constitutional Carry - Removed requirement for a permit to carry a concealed firearm for eligible individuals. 2023 (L): Expanded Constitutional Carry - Allowed permitless carry in more locations, including schools and college campuses. |
| Georgia | 2006 (L): Stand Your Ground Law - Removed the duty to retreat before using force in self-defense situations. 2008 (L): Public Transportation Carry - Allowed concealed carry permit holders to carry firearms on public transportation. 2010 (L): Bar and Restaurant Carry - Permitted carrying firearms in bars and restaurants that serve alcohol. 2014 (L): Guns Everywhere Act - Expanded where guns could be carried, including some government buildings, bars, and churches (with permission). 2016 (L): Campus Carry - Allowed concealed carry permit holders to carry firearms on public college campuses. 2017 (L): Courthouse Carry - Allowed firearms in certain areas of courthouses. 2022 (L): Constitutional Carry - Removed requirement for a permit to carry a concealed firearm for eligible individuals. 2022 (L): Second Amendment Protection Act - Prohibited state and local law enforcement from enforcing certain federal gun regulations. 2023 (L): Campus Carry Expansion - Extended campus carry rights to include more areas of college campuses. |
| Hawaii | 2006 (S): Electric Gun Ban - Prohibited the possession, sale, or transfer of electric guns (e.g., stun guns and tasers). 2013 (S): Registration of Firearms - Required registration of firearms brought into the state within 3 days of arrival. 2016 (S): Firearms Owners Registration - Required gun owners to be entered into an FBI database for continuous criminal record monitoring. 2018 (S): Bump Stock Ban - Prohibited the manufacture, import, sale, transfer, and possession of bump stocks and other rapid-fire devices. 2019 (S): Gun Violence Protective Orders - Established a red flag law allowing courts to temporarily remove firearms from individuals deemed a risk. 2020 (S): Ghost Gun Regulation - Required ghost guns to have serial numbers and be registered. 2020 (S): Ammunition Purchase Background Checks - Required background checks for ammunition purchases. 2022 (S): Concealed Carry Restrictions - Implemented strict requirements for obtaining a concealed carry permit in response to the NYSRPA v. Bruen decision. 2023 (S): Assault Weapons Ban Enhancement - Strengthened existing assault weapons regulations. |
| Idaho | 2006 (L): Stand Your Ground Law - Removed the duty to retreat before using force in self-defense situations. 2008 (L): Preemption Law Enhancement - Strengthened state preemption of firearms laws, limiting local governments' ability to regulate firearms. 2014 (L): Guns on Campus - Allowed concealed carry permit holders to carry firearms on public college and university campuses. 2015 (L): Constitutional Carry - Allowed permitless carry of concealed firearms for residents 21 and older. 2016 (L): Constitutional Carry Expansion - Extended permitless carry rights to non-residents. 2018 (L): Stand Your Ground Enhancement - Expanded legal protections for use of force in self-defense situations. 2019 (L): Lowered Concealed Carry Age - Reduced the minimum age for concealed carry without a permit from 21 to 18 for residents. 2020 (L): Second Amendment Sanctuary State - Prohibited state officials from enforcing federal gun laws deemed to violate the Second Amendment. 2021 (L): Emergency Powers Limitation - Prohibited government officials from restricting gun rights during emergencies or disasters. 2023 (L): Campus Carry Enhancement - Expanded areas on college campuses where firearms can be carried. |
| Illinois | 2013 (L): Concealed Carry Act - Allowed concealed carry of firearms, making Illinois the last state to do so. 2013 (S): Firearm Concealed Carry Act - Established strict requirements for obtaining a concealed carry license. 2014 (S): Private Sale Background Check Law - Required private sellers to verify buyer's FOID card with state police. 2018 (S): 72-Hour Waiting Period Law - Enacted a 72-hour waiting period for all firearm purchases. 2019 (S): Firearm Dealer License Certification Act - Required gun dealers to obtain a state license in addition to federal license. 2019 (S): Red Flag Law - Allowed family or law enforcement to petition for temporary firearm removal from at-risk individuals. 2021 (S): Universal Background Check Expansion - Extended background check requirements to all private firearm transfers. 2022 (S): Ghost Gun Ban - Prohibited the sale and manufacture of unserialized and untraceable firearms. 2023 (S): Protect Illinois Communities Act - Banned sale, manufacture, and possession of assault weapons and high-capacity magazines. 2024 (S): FOID Fingerprint Requirement - Mandated fingerprint submission for FOID card applicants. |
| Indiana | 2011 (L): Indiana passed a law prohibiting private employers from banning firearms in locked vehicles on company property. 2014 (L): The state enacted a law allowing guns in school parking lots if locked in a vehicle. 2019 (L): Indiana expanded its "stand your ground" law to provide civil immunity for justified use of force. 2020 (L): The state extended the duration of handgun licenses from four years to five years. 2021 (L): Indiana eliminated the fee for lifetime carry permits. 2022 (L): The state adopted permitless carry, allowing most adults to carry a handgun without a license. 2023 (S): Indiana passed a law requiring safe storage of firearms in homes with children under 18 years old. 2023 (S): The state enacted a law prohibiting the sale of firearms to individuals under protective orders for domestic violence. 2024 (L): Indiana expanded legal protections for armed citizens who use force in self-defense situations. 2024 (S): The state implemented a law requiring background checks for private party firearm transfers at gun shows. |
| Iowa | 2010 (L): Shall Issue Law - Required sheriffs to issue concealed carry permits to qualified applicants. 2011 (L): Recreational Vehicle Carry Law - Removed prohibition on carrying firearms while operating snowmobiles or ATVs. 2017 (L): Stand Your Ground Law - Implemented stand-your-ground provisions and allowed supervised handgun use by children under 14. 2021 (L): Permitless Carry Law - Allowed adults to carry concealed firearms without a permit. 2021 (L): Firearms Preemption Law - Prevented local governments from implementing stricter firearms restrictions than state law. 2021 (S): Domestic Violence Prohibition - Banned firearms sales to individuals subject to certain protective orders or misdemeanor convictions. 2022 (L): Constitutional Amendment - Added strict scrutiny standard for gun laws to state constitution. 2023 (L): Self-Defense Protection Expansion - Increased legal protections for gun owners in self-defense situations. 2024 (S): Gun Show Background Check Law - Required background checks for private party firearm transfers at gun shows. 2024 (L): Concealed Carry Location Expansion - Allowed concealed carry in more locations, including some government buildings. |
| Kansas | 2006 (L): Shall Issue Law - Required issuance of concealed carry permits to qualified applicants. 2013 (L): Second Amendment Protection Act - Declared federal laws regulating firearms within Kansas unenforceable. 2014 (L): Public Building Carry Law - Allowed carrying firearms in public buildings lacking adequate security measures. 2015 (L): Constitutional Carry Law - Permitted individuals to carry concealed firearms without a permit. 2017 (L): Campus Carry Law - Prohibited state universities from banning concealed firearms on campus. 2018 (S): Domestic Violence Prohibition - Banned individuals convicted of misdemeanor domestic violence from possessing firearms. 2021 (L): Concealed Carry Age Reduction - Lowered minimum age for concealed carry from 21 to 18 with training. 2022 (L): Out-of-State Permit Recognition - Recognized all out-of-state concealed carry permits. 2023 (L): Self-Defense Protection Expansion - Increased legal protections for armed citizens using force in self-defense. 2024 (S): Safe Storage Law - Required safe storage of firearms in homes with minors. |
| Kentucky | 2013 (L): Senate Bill 150 - Allowed concealed carry of firearms in vehicles without a permit. 2014 (L): House Bill 128 - Permitted local governments to allow concealed carry in government buildings. 2016 (L): Senate Bill 14 - Allowed active duty military members aged 18-20 to obtain concealed carry permits. 2017 (L): House Bill 417 - Recognized concealed carry permits from other states. 2019 (L): Constitutional Carry Law - Allowed permitless concealed carry for individuals 21 and older who are legally eligible to possess a firearm. 2020 (L): Senate Bill 15 - Allowed concealed carry in schools by staff members serving as school security officers. 2021 (L): House Bill 236 - Prohibited state and local law enforcement from enforcing federal firearms bans. 2022 (L): House Bill 29 - Eliminated location restrictions for concealed carry permit holders, including in bars and other businesses serving alcohol. 2023 (L): Senate Bill 150 - Strengthened preemption laws, preventing local governments from enacting their own firearm regulations. |
| Louisiana | 2006 (L): "Stand Your Ground" Law - Expanded self-defense protections, including no duty to retreat. 2010 (L): Restaurant Carry Law - Allowed concealed carry permit holders to bring firearms into restaurants that serve alcohol. 2012 (L): Right to Bear Arms Constitutional Amendment - Strengthened right to keep and bear arms, requiring strict scrutiny for gun regulations. 2013 (L): Concealed Carry Privacy Act - Prohibited release of concealed carry permit holder information to the public. 2014 (L): Lifetime Concealed Carry Permit Law - Allowed lifetime concealed carry permits. 2015 (L): Firearms in Alcohol-Serving Establishments Law - Reduced penalties for carrying a firearm in an alcohol-serving establishment with a concealed carry permit. 2017 (L): Military Personnel Concealed Carry Expansion - Expanded concealed carry rights for military personnel. 2019 (L): Church Carry Law - Allowed concealed carry in places of worship with permission from religious leaders. 2021 (L): Concealed Carry Disclosure Law - Removed requirement for concealed carry permit holders to disclose their permit status to law enforcement. 2022 (L): Louisiana Firearms Protection Act - Prohibited state and local enforcement of federal gun restrictions enacted after January 1, 2021. 2023 (L): Constitutional Carry Law - Allowed permitless concealed carry for individuals 18 and older who are legally able to possess a firearm. |
| Maine | 2003 (L): Concealed Carry Reciprocity Law - Recognized concealed carry permits from certain other states. 2011 (L): "Guns in Cars" Law - Allowed employees to keep firearms in their locked vehicles at work. 2015 (L): Constitutional Carry Law - Permitted permitless concealed carry for residents 21 and older (18 for military members). 2015 (S): Dating Partner Gun Ban - Extended firearm possession restrictions to include individuals subject to protective orders for dating partners. 2019 (S): "Yellow Flag" Law - Allowed courts to order temporary removal of firearms from individuals deemed to be at risk of harming themselves or others. 2019 (L): Hunting with Suppressors Law - Legalized the use of noise suppressors while hunting. 2021 (L): Second Amendment Sanctuary Law - Prohibited state and local resources from being used to enforce certain federal gun regulations. 2023 (S): Safe Storage Law - Required firearms to be securely stored when not in use to prevent unauthorized access. 2023 (S): Waiting Period Law - Implemented a 72-hour waiting period for firearm purchases. 2023 (S): Background Check Expansion - Required background checks for private gun sales and transfers, with some exceptions. |
| Maryland | 2002 (S): Integrated Ballistics Identification System Law - Required handgun manufacturers to provide shell casings for all handguns sold in the state. 2013 (S): Firearm Safety Act - Banned assault weapons, limited magazine capacity to 10 rounds, and required fingerprinting for handgun purchases. 2015 (S): Domestic Violence Gun Surrender Law - Required individuals convicted of certain domestic violence crimes to surrender firearms. 2018 (S): Red Flag Law - Allowed courts to temporarily remove firearms from individuals deemed to be a risk to themselves or others. 2021 (S): Ghost Gun Ban - Prohibited the sale, receipt, and transfer of unfinished frames and receivers without serial numbers. 2022 (S): Handgun Qualification License Study - Required a study on the effectiveness of the Handgun Qualification License requirement. 2023 (S): Gun Safety Act - Prohibited carrying firearms in "sensitive places" like schools and government buildings. 2023 (S): Bruen Compliance Act - Revised concealed carry permit process in response to the Supreme Court's Bruen decision, but maintained strict requirements. 2023 (S): Raise the Age Act - Increased minimum age to purchase rifles and shotguns from 18 to 21, with some exceptions. 2024 (S): Firearm Industry Accountability Act - Allowed lawsuits against gun manufacturers and dealers for certain gun crimes. |
| Massachusetts | 2004 (S): Assault Weapons Ban Renewal - Renewed and strengthened the state's ban on assault weapons after the federal ban expired. 2014 (S): Gun Violence Reduction Act - Expanded background checks, gave police chiefs more discretion in issuing firearms licenses, and required schools to develop plans for active shooters. 2017 (S): Bump Stock Ban - Prohibited the possession of bump stocks and other rapid-fire devices. 2018 (S): Extreme Risk Protection Order Law - Allowed family members or law enforcement to petition courts to temporarily remove firearms from individuals deemed to be a risk. 2018 (S): Urban Gun Violence Prevention Bill - Established a center for gun violence prevention and funded community-based programs. 2020 (S): Ghost Gun Regulation - Required serialization of "ghost guns" and banned 3D-printed firearms. 2022 (L): Bruen Decision Implementation - Required adjustments to concealed carry licensing process due to the Supreme Court's Bruen decision. 2023 (S): Firearm Data Reporting Act - Required more detailed reporting of firearm sales and transfers data. 2024 (S): Safe Storage Law Enhancement - Strengthened requirements for secure storage of firearms when not in use. 2024 (S): Gun Dealer Regulation Act - Implemented stricter oversight and security requirements for firearms dealers. |
| Michigan | 2001 (L): Shall-Issue Concealed Carry Law - Changed Michigan from a "may-issue" to a "shall-issue" state for concealed carry permits. 2006 (L): Self-Defense Act - Expanded self-defense rights, including a "stand your ground" provision. 2012 (L): Emergency Powers Limitation Act - Prohibited the governor from restricting firearm possession during states of emergency. 2015 (L): Concealed Pistol License Streamlining Act - Shifted the responsibility for issuing concealed carry permits from county gun boards to county clerks. 2017 (L): Air Gun Reclassification Law - Removed air guns from the definition of firearms. 2020 (L): Capitol Carry Clarification - Affirmed the right to openly carry firearms in the state Capitol building. 2023 (S): Safe Storage Law - Required gun owners to securely store firearms if minors are present in the home. 2023 (S): Universal Background Check Law - Extended background check requirements to all firearm sales, including private transactions. 2023 (S): Extreme Risk Protection Order Act - Implemented a "red flag" law allowing courts to temporarily remove firearms from individuals deemed a risk. 2024 (S): Domestic Violence Firearm Restriction - Prohibited individuals convicted of domestic violence misdemeanors from possessing firearms for a specified period. |
| Minnesota | 2003 (L): Minnesota Citizens' Personal Protection Act - Established a "shall-issue" system for concealed carry permits. 2005 (L): Stand Your Ground Law - Expanded self-defense rights, removing the duty to retreat in certain situations. 2014 (S): Domestic Violence Gun Ban - Prohibited individuals convicted of domestic violence or subject to protective orders from possessing firearms. 2015 (L): Suppressor Legalization - Legalized the ownership and use of firearm suppressors. 2019 (S): Gun Violence Protective Order Law - Allowed family members and law enforcement to petition courts for temporary removal of firearms from individuals deemed a risk. 2021 (L): Capitol Carry Notification Law - Required the Department of Public Safety to notify permit holders about their right to carry in the State Capitol complex. 2023 (S): Universal Background Check Law - Required background checks for private firearm transfers, with some exceptions. 2023 (S): Red Flag Law - Implemented an Extreme Risk Protection Order system, allowing courts to temporarily remove firearms from individuals deemed a significant danger. 2023 (S): Safe Storage Law - Required firearms to be securely stored to prevent unauthorized access, especially by minors. 2024 (S): Ghost Gun Regulation - Restricted the sale and possession of unserialized firearms and unfinished frames or receivers. |
| Mississippi | 2010 (L): Enhanced Carry Permit Law - Created an enhanced concealed carry permit allowing carry in some previously restricted locations. 2011 (L): Castle Doctrine Expansion - Expanded self-defense protections to include places of business and vehicles. 2013 (L): Firearms Protection Act - Prohibited state enforcement of any future federal gun laws that might conflict with the state constitution. 2014 (L): Second Amendment Sales Tax Holiday - Established an annual tax-free weekend for firearms, ammunition, and hunting supplies. 2015 (L): Church Protection Act - Allowed churches to designate members to carry firearms for security purposes. 2016 (L): Constitutional Carry Law - Permitted permitless concealed carry of firearms for those legally allowed to possess a firearm. 2018 (L): School Safety Act - Allowed school employees with enhanced carry permits to carry firearms on school grounds. 2021 (L): Second Amendment Preservation Act - Prohibited state agencies from enforcing certain federal gun regulations. 2022 (L): Permit-less Carry Age Reduction - Lowered the age for permit-less carry from 21 to 18. 2023 (L): Home-Based Firearm Business Protection Act - Prevented local governments from restricting home-based firearm businesses. |
| Missouri | 2003 (L): Concealed Carry Law - Allowed concealed carry of firearms with a permit. 2007 (L): Castle Doctrine Law - Expanded self-defense rights, removing duty to retreat in one's home or vehicle. 2014 (L): Open Carry Protection Act - Prohibited local governments from banning open carry for permit holders. 2016 (L): Constitutional Carry Law - Allowed permitless concealed carry for those legally able to possess firearms. 2016 (L): Stand Your Ground Law - Removed duty to retreat before using force in self-defense in any location. 2017 (L): Campus Carry Law - Allowed concealed carry on college campuses and in most areas of the state capitol. 2019 (S): Domestic Violence Gun Restriction - Prohibited firearm possession for those convicted of domestic violence misdemeanors. 2021 (L): Second Amendment Preservation Act - Declared certain federal gun laws "invalid" in Missouri and prohibited state law enforcement from enforcing them. 2022 (L): No-Knock Warrant Restriction - Limited no-knock warrants, potentially affecting firearm-related searches. 2023 (L): Armed Teacher Protection Act - Provided legal protections for teachers who carry firearms in schools with district permission. |
| Montana | 2009 (L): Montana Firearms Freedom Act - Declared that firearms made and kept in Montana are not subject to federal regulation. 2011 (L): Right to Hunt Amendment - Added the right to hunt, fish, and trap to the state constitution. 2015 (L): Gun Show Loophole Closure Prevention Act - Prohibited local governments from requiring background checks at gun shows. 2016 (L): Permitless Carry Law - Allowed permitless concealed carry in most areas outside of city limits. 2019 (L): Campus Carry Law - Allowed concealed carry on college campuses for permit holders. 2021 (L): Constitutional Carry Act - Expanded permitless concealed carry to most public places, including within city limits. 2021 (L): Montana Federal Firearm, Magazine, and Ammunition Ban Enforcement Prohibition Act - Prohibited state enforcement of certain federal gun laws. 2021 (L): Home-Owned Firearms Protection Act - Prohibited financial institutions from discriminating against firearms businesses. 2023 (L): Second Amendment Sanctuary State Act - Declared Montana a "sanctuary state" for gun rights, further limiting enforcement of federal gun laws. 2024 (L): Montana First Act - Gave preference to in-state firearm and ammunition manufacturers for state law enforcement contracts. |
| Nebraska | 2006 (L): Concealed Handgun Permit Act - Established a shall-issue concealed carry permit system. 2009 (L): Castle Doctrine Law - Expanded self-defense rights, removing duty to retreat in one's home. 2010 (L): Firearms in Vehicles Law - Allowed transportation of unloaded firearms in vehicles without a permit. 2011 (L): Preemption Law - Prohibited local governments from enacting firearm regulations more restrictive than state law. 2015 (L): Gun Rights Restoration Act - Created a process for restoring gun rights to certain individuals with past felony convictions. 2018 (S): Extreme Risk Protection Order Act - Allowed family members or law enforcement to petition courts for temporary removal of firearms from individuals deemed a risk. 2020 (L): First Responder Concealed Carry Act - Allowed off-duty first responders to carry concealed firearms in more locations. 2021 (L): Second Amendment Preservation Act - Prohibited state enforcement of certain federal gun laws deemed to infringe on the Second Amendment. 2023 (L): Constitutional Carry Law - Allowed permitless concealed carry for individuals 21 and older who can legally possess a firearm. 2024 (S): Safe Storage Act - Required firearms to be securely stored when not in use to prevent unauthorized access. |
| Nevada | 2007 (L): Castle Doctrine Law - Expanded self-defense rights, removing duty to retreat in one's home. 2011 (L): State Preemption Law - Strengthened state preemption of local firearm regulations. 2015 (L): Campus Carry Law - Allowed concealed carry permit holders to store firearms in locked vehicles on college campuses. 2015 (S): Domestic Violence Gun Ban - Prohibited individuals convicted of domestic violence misdemeanors from possessing firearms. 2016 (S): Background Check Initiative - Passed a ballot measure requiring background checks for private gun sales (implementation delayed until 2020). 2017 (S): "Red Flag" Law - Allowed family members or law enforcement to petition courts for temporary removal of firearms from individuals deemed a risk. 2019 (S): Omnibus Gun Bill - Implemented storage requirements, banned bump stocks, and lowered blood alcohol limit for firearm possession. 2020 (S): Background Check Law Implementation - Officially implemented the 2016 background check initiative for private gun sales. 2021 (S): Ghost Gun Ban - Prohibited the sale and possession of unserialized firearms. 2023 (S): Assault Weapons Restrictions - Implemented stricter regulations on semi-automatic firearms classified as assault weapons. |
| New Hampshire | 2003 (L): Castle Doctrine Law - Expanded self-defense rights, removing duty to retreat in one's home. 2006 (L): Preemption Law - Strengthened state preemption of local firearm regulations. 2011 (L): Stand Your Ground Law - Removed duty to retreat before using deadly force in self-defense in any place one has a right to be. 2015 (L): Knife Rights Law - Repealed bans on switchblades, dirks, and stilettos, aligning knife laws with firearm laws. 2017 (L): Constitutional Carry Law - Allowed permitless concealed carry for anyone who can legally possess a firearm. 2018 (S): Domestic Violence Gun Restrictions - Prohibited gun possession by individuals subject to domestic violence protective orders. 2019 (S): Gun-Free School Zones Act - Prohibited carrying firearms on school property, with exceptions for law enforcement and authorized individuals. 2020 (L): Second Amendment Sanctuary Towns - Several towns declared themselves "Second Amendment sanctuaries," though these had no legal effect at the state level. 2022 (L): State Preemption Enhancement - Further strengthened state preemption, explicitly prohibiting local governments from regulating firearms or ammunition. 2023 (S): Safe Storage Law - Required firearms to be securely stored when not under direct control of the owner, particularly in homes with children. |
| New Jersey | 2002 (S): Childproof Handgun Law - Required smart gun technology once commercially available. 2013 (S): Gun Violence Prevention Package - Included multiple measures such as banning armor-piercing bullets and increasing penalties for gun trafficking. 2018 (S): Magazine Capacity Reduction - Reduced maximum magazine capacity from 15 to 10 rounds. 2018 (S): Extreme Risk Protective Order Act - Implemented a "red flag" law allowing temporary removal of firearms from at-risk individuals. 2022 (S): Concealed Carry Restriction Law - Implemented strict requirements for concealed carry in response to the Supreme Court's Bruen decision. |
| New Mexico | 2013 (S): Background Check Expansion - Required background checks at gun shows. 2019 (S): Universal Background Check Law - Mandated background checks for nearly all firearm transfers. 2020 (S): Extreme Risk Firearm Protection Order Act - Allowed courts to temporarily remove firearms from at-risk individuals. 2023 (S): Firearm Safe Storage Act - Required secure storage of firearms when not in use. 2023 (S): Assault Weapon Ban - Prohibited the sale and possession of certain semi-automatic firearms defined as assault weapons. |
| New York | 2013 (S): NY SAFE Act - Expanded assault weapons ban, limited magazine capacity, and required background checks for ammunition purchases. 2019 (S): Red Flag Law - Allowed courts to order temporary removal of firearms from individuals deemed dangerous. 2020 (S): Ghost Gun Ban - Prohibited the possession and sale of ghost guns and unfinished receivers. 2022 (S): Concealed Carry Improvement Act - Implemented strict requirements for concealed carry permits in response to the Supreme Court's Bruen decision. 2023 (S): Ammunition Background Check Implementation - Finally implemented the ammunition background check system from the 2013 SAFE Act. |
| North Carolina | 2011 (L): Castle Doctrine Expansion - Strengthened self-defense rights in homes, workplaces, and vehicles. 2013 (L): Gun Rights and Privacy Act - Prohibited destruction of surrendered firearms and allowed concealed carry in more locations. 2015 (L): Firearms Freedom Act - Removed some restrictions on short-barreled rifles and eased rules for suppressors. 2015 (S): Domestic Violence Gun Surrender Law - Required domestic abusers subject to protective orders to surrender firearms. 2017 (L): Permitless Concealed Carry for Military Veterans - Allowed veterans to carry concealed without a permit. 2019 (S): Extreme Risk Protection Order Act - Proposed "red flag" law (not passed, but significant in state gun policy debate). 2020 (L): Second Amendment Protection Act - Allowed gun stores to remain open during states of emergency. 2021 (L): Church Carry Law - Allowed concealed carry permit holders to carry firearms in churches that also serve as schools outside of school hours. 2023 (L): Pistol Purchase Permit Repeal - Eliminated the requirement for a permit to purchase a handgun. 2024 (S): Safe Storage Awareness Campaign - Implemented a statewide campaign to promote safe firearm storage (a compromise measure after stricter proposals). |
| North Dakota | 2007 (L): Castle Doctrine Law - Expanded self-defense rights, removing duty to retreat in one's home. 2013 (L): Firearms Freedom Act - Declared that firearms made and kept in North Dakota are not subject to federal regulation. 2015 (L): Constitutional Carry Law - Allowed permitless concealed carry for residents. 2017 (L): Campus Carry Law - Allowed concealed firearms in vehicles on school property. 2019 (L): Armed First Responder Program - Allowed armed first responders in schools. 2021 (L): Second Amendment Sanctuary State Law - Prohibited state agencies from enforcing federal gun laws deemed to infringe on gun rights. 2023 (L): Public Gathering Law Repeal - Removed restrictions on carrying firearms at public gatherings. |
| Ohio | 2004 (L): Concealed Carry Law - Established shall-issue concealed carry permits. 2008 (L): Castle Doctrine Law - Expanded self-defense rights in homes and vehicles. 2011 (L): Restaurant Carry Law - Allowed concealed carry in establishments serving alcohol. 2017 (L): Campus Carry Law - Allowed colleges to decide whether to permit concealed carry on campus. 2020 (L): Stand Your Ground Law - Removed duty to retreat before using force in self-defense. 2022 (L): Constitutional Carry Law - Allowed permitless concealed carry for those 21 and older who can legally possess a firearm. 2023 (L): Armed Teacher Training Reduction Act - Reduced required training hours for school staff to carry firearms. 2024 (S): Extreme Risk Protection Order Act - Implemented a "red flag" law allowing temporary removal of firearms from at-risk individuals (notable as a rare restrictive measure in Ohio). |
| Oklahoma | 2012 (L): Open Carry Law - Allowed licensed individuals to openly carry handguns in public. 2015 (L): Expanded Stand Your Ground - Extended "Stand Your Ground" protections to places of worship. 2019 (L): Constitutional Carry - Allowed permitless carry of firearms for individuals 21 and older (18 for military members). 2020 (L): Anti-Red Flag Law - Prohibited the state from enforcing federal red flag laws. 2021 (L): Second Amendment Sanctuary State - Declared Oklahoma a Second Amendment Sanctuary, prohibiting state agencies from enforcing certain federal gun laws. 2022 (L): Firearms Industry Nondiscrimination Act - Prohibited financial institutions from discriminating against firearm businesses. 2023 (L): School Marshal Program - Allowed school employees to carry firearms on school grounds after completing training. |
| Oregon | 2001 (S): Background Check Expansion - Required background checks for all firearm transfers at gun shows. 2015 (S): Universal Background Checks - Mandated background checks for private gun sales and transfers. 2017 (S): Extreme Risk Protection Order - Implemented "red flag" law allowing courts to temporarily remove firearms from individuals deemed a risk. 2018 (S): Domestic Violence Prohibition - Expanded firearm prohibition to include intimate partners convicted of domestic violence or under restraining orders. 2021 (S): Safe Storage Law - Required gun owners to securely store firearms when not in use. 2022 (S): Ghost Gun Ban - Prohibited the manufacture, sale, and possession of unserialized firearms ("ghost guns"). 2023 (S): Measure 114 - Implemented permit-to-purchase requirement, safety training, and banned magazines over 10 rounds (Note: Implementation was delayed due to legal challenges). 2024 (S): Age Restriction - Raised minimum age to purchase firearms from 18 to 21 (with exceptions for military and law enforcement). |
| Pennsylvania | 2005 (L): Castle Doctrine Expansion - Expanded self-defense rights in one's home and vehicle. 2011 (L): Stand Your Ground Law - Removed duty to retreat before using deadly force in self-defense situations outside the home. 2014 (S): Act 80 - Required individuals subject to protection from abuse orders to surrender their firearms more quickly. 2018 (S): Act 79 - Strengthened firearm relinquishment requirements for domestic abusers. 2019 (L): Preemption Reinforcement - Strengthened state preemption of local firearm regulations. 2020 (S): Background Check Expansion - Extended background check requirements to all firearms, including long guns. 2023 (S): Extreme Risk Protection Order - Implemented "red flag" law allowing temporary removal of firearms from individuals deemed at risk (Note: This was a significant change, but implementation may have faced legal challenges). |
| Rhode Island | 2005 (S): Domestic Violence Prohibition - Prohibited individuals convicted of domestic violence misdemeanors from possessing firearms. 2012 (S): Background Check Enhancement - Required background checks for all employees of gun dealers. 2018 (S): Red Flag Law - Implemented Extreme Risk Protection Orders, allowing temporary removal of firearms from individuals deemed a risk. 2019 (S): Ghost Gun Ban - Prohibited the manufacture, sale, and possession of 3D-printed firearms and other untraceable "ghost guns." 2021 (S): Straw Purchase Prevention - Prohibited purchasing a firearm on behalf of someone else who is legally prohibited from owning one. 2022 (S): High-Capacity Magazine Ban - Prohibited possession, manufacture, or sale of magazines capable of holding more than 10 rounds. 2022 (S): Age Restriction - Raised minimum age to purchase firearms and ammunition from 18 to 21 (with exceptions for law enforcement and military). 2023 (S): Assault Weapons Ban - Prohibited the sale, transfer, and possession of certain semi-automatic firearms defined as "assault weapons." |
| South Carolina | 2006 (L): Protection of Persons and Property Act - Expanded Castle Doctrine and removed duty to retreat in self-defense situations. 2014 (L): Concealed Carry in Restaurants - Allowed concealed carry permit holders to bring firearms into establishments serving alcohol, provided they don't consume alcohol. 2016 (L): Reciprocity Expansion - Recognized concealed weapon permits from all other states. 2017 (L): Church Carry - Allowed concealed carry in churches with permission from church leadership. 2021 (L): Open Carry with Training Act - Allowed open carry of handguns for individuals with concealed weapon permits. 2022 (L): Constitutional Carry - Passed law allowing permitless carry of firearms, both open and concealed (Note: This was vetoed by the governor but may have been overridden or reintroduced later). 2023 (L): Second Amendment Sanctuary Act - Prohibited state and local law enforcement from enforcing certain federal gun restrictions. |
| South Dakota | 2005 (L): Stand Your Ground Law - Removed duty to retreat before using deadly force in self-defense situations. 2009 (L): Firearms Freedom Act - Declared that firearms manufactured and retained in South Dakota are not subject to federal regulation. 2015 (L): Enhanced Carry Permit - Created an optional enhanced concealed carry permit allowing carry in more locations. 2017 (L): Capitol Carry - Allowed concealed carry in the state capitol building for enhanced permit holders. 2019 (L): Constitutional Carry - Implemented permitless carry, allowing legal gun owners to carry concealed without a permit. 2022 (L): Second Amendment Sanctuary - Declared South Dakota a Second Amendment sanctuary state, limiting enforcement of certain federal gun laws. 2023 (L): Strengthened Preemption - Reinforced state preemption of local firearm regulations, preventing cities from enacting stricter gun laws. 2024 (L): Campus Carry - Allowed concealed carry on public university campuses for permit holders (Note: If this hasn't been enacted yet, it was a significant proposal during this period). |
| Tennessee | 2009 (L): Guns in Bars - Allowed handgun carry permit holders to bring firearms into restaurants serving alcohol. 2010 (L): Guns in Parks - Allowed handgun carry permit holders to carry in state and local parks. 2013 (L): Guns in Parking Lots - Prohibited employers from banning firearms in locked vehicles on company property. 2015 (L): Lifetime Carry Permits - Introduced lifetime handgun carry permits. 2016 (L): Campus Carry - Allowed full-time faculty and staff at public colleges to carry handguns on campus. 2017 (L): Enhanced Handgun Carry Permit - Created a new permit with fewer restrictions than the standard permit. 2020 (L): Constitutional Carry - Implemented permitless carry for adults 21 and older. 2021 (L): Second Amendment Sanctuary - Declared Tennessee a Second Amendment sanctuary state, limiting enforcement of certain federal gun laws. 2022 (L): Permitless Open Carry for 18-20 - Extended permitless carry rights to individuals aged 18-20 for open carry. 2023 (L): Armed School Staff - Allowed some school staff to carry firearms on school grounds after completing training. |
| Texas | 2007 (L): Castle Doctrine Expansion - Expanded self-defense rights and removed duty to retreat in one's home, vehicle, or workplace. 2011 (L): Parking Lot Protection - Prohibited employers from banning firearms in locked vehicles on company property. 2015 (L): Open Carry - Allowed licensed handgun owners to openly carry handguns in most public places. 2016 (L): Campus Carry - Permitted concealed carry of handguns on public university campuses. 2019 (L): Expanded Carry Locations - Allowed licensed carry in places of worship and removed some restrictions on school carry. 2021 (L): Constitutional Carry - Implemented permitless carry, allowing legal gun owners to carry handguns without a license. 2021 (L): Second Amendment Sanctuary - Declared Texas a Second Amendment sanctuary state, prohibiting state agencies from enforcing certain federal gun laws. 2023 (L): Hotel Carry Protection - Prohibited hotels from restricting guests from storing firearms in their rooms. 2023 (L): Lower Age for Handgun Purchases - Allowed 18-20 year-olds to obtain handgun carry licenses with military service or honorable discharge. |
| Utah | 2004 (L): Preemption Law - Strengthened state preemption of local firearm regulations, preventing cities from enacting stricter gun laws. 2006 (L): Concealed Carry on School Grounds - Allowed concealed carry permit holders to carry firearms on school grounds. 2010 (L): Constitutional Carry for Unloaded Firearms - Allowed open or concealed carry of unloaded firearms without a permit. 2013 (L): Domestic Violence Gun Surrender - Required individuals subject to protective orders to surrender firearms (Note: This is labeled L because it was less restrictive than federal law). 2015 (L): Campus Carry Protection - Prohibited public colleges from creating "gun-free zones" on campus. 2021 (L): Constitutional Carry - Implemented permitless carry, allowing legal gun owners to carry concealed firearms without a permit. 2021 (L): Second Amendment Sanctuary - Declared Utah a Second Amendment sanctuary state, limiting enforcement of certain federal gun laws. 2023 (L): Strengthened Preemption - Further reinforced state preemption, imposing penalties on local governments that enact gun regulations. 2024 (L): School Guardian Program - Allowed trained school employees to carry firearms on school grounds (Note: If not yet enacted, this was a significant proposal during this period). |
| Vermont | 2018 (S): S.55 Gun Control Package - This was a significant shift in Vermont's traditionally permissive gun laws, including:  Universal Background Checks: Required background checks for private gun sales. Age Restriction: Raised minimum age to purchase firearms to 21 (with exceptions). Magazine Capacity Limit: Banned magazines over 10 rounds for long guns and 15 for handguns. Bump Stock Ban: Prohibited possession and use of bump stocks.  2018 (S): H.422 Domestic Violence Law - Allowed police to confiscate firearms from those accused of domestic violence. 2019 (S): Waiting Period Law - Implemented a 24-hour waiting period for handgun purchases. 2022 (S): Ghost Gun Regulation - Prohibited the manufacture, sale, and possession of unserialized firearms ("ghost guns"). 2023 (S): Safe Storage Law - Required firearms to be securely stored when not in use to prevent unauthorized access. |
| Virginia | 2004 (L): Concealed Carry Reform - Streamlined the concealed carry permit process and expanded reciprocity with other states. 2010 (L): Restaurant Carry - Allowed concealed carry permit holders to bring firearms into restaurants serving alcohol, provided they don't consume alcohol. 2012 (L): One-Gun-a-Month Repeal - Removed the law limiting handgun purchases to one per month. 2016 (L): Domestic Violence Gun Rights - Restored gun rights to individuals convicted of certain domestic violence misdemeanors after a waiting period. 2020 (S): Gun Control Package - Implemented several new restrictions:  Universal Background Checks Red Flag Law (Extreme Risk Protection Orders) One-Gun-a-Month Rule Reinstatement Reporting of Lost or Stolen Firearms  2021 (S): Ghost Gun Ban - Prohibited the manufacture, sale, and possession of unserialized firearms ("ghost guns"). 2022 (S): Safe Storage Law - Required firearms to be securely stored in homes with minors present. 2023 (L): School Guardian Program - Allowed trained school staff to carry firearms on school grounds (Note: If not enacted, this was a significant proposal during this period). |
| Washington | 2014 (S): Universal Background Checks - Required background checks for all firearm transfers, including private sales (Initiative 594). 2016 (S): Extreme Risk Protection Orders - Implemented "red flag" law allowing temporary removal of firearms from individuals deemed a risk (Initiative 1491). 2018 (S): Age Restrictions and Safe Storage - Raised minimum age to purchase semi-automatic rifles to 21 and implemented safe storage requirements (Initiative 1639). 2019 (S): Domestic Violence Gun Surrender - Strengthened laws requiring surrender of firearms by individuals subject to certain protective orders. 2020 (S): Ghost Gun Regulations - Restricted manufacture and sale of untraceable firearms and undetectable firearms. 2021 (S): Open Carry Restriction - Prohibited open carry of firearms at or near permitted demonstrations and at the state capitol. 2022 (S): High-Capacity Magazine Ban - Prohibited the manufacture, sale, and distribution of magazines with a capacity of more than 10 rounds. 2023 (S): Assault Weapons Ban - Prohibited the sale, manufacture, and import of certain semi-automatic firearms defined as "assault weapons" (Note: If not enacted, this was a significant proposal during this period). |
| West Virginia | 2004 (L): Castle Doctrine - Expanded self-defense rights in one's home and removed duty to retreat. 2008 (L): Parking Lot Protection - Prohibited employers from banning firearms in locked vehicles on company property. 2015 (L): Municipal Gun Law Preemption - Strengthened state preemption of local firearm regulations. 2016 (L): Constitutional Carry - Implemented permitless carry, allowing legal gun owners to carry concealed firearms without a permit. 2018 (L): Campus Carry - Allowed concealed carry on college campuses (with some restrictions). 2020 (L): Second Amendment Preservation Act - Prohibited state agencies from enforcing certain federal gun control laws. 2021 (L): Expanded Concealed Carry Locations - Allowed concealed carry in more public places, including some government buildings. 2022 (L): Hunting with Suppressors - Legalized the use of suppressors for hunting. 2023 (L): School Guardian Program - Allowed trained school staff to carry firearms on school grounds (Note: If not enacted, this was a significant proposal during this period). |
| Wisconsin | 2011 (L): Concealed Carry - Legalized concealed carry of firearms with a permit. 2011 (L): Castle Doctrine - Expanded self-defense rights in one's home, vehicle, and place of business. 2015 (L): Waiting Period Repeal - Eliminated the 48-hour waiting period for handgun purchases. 2015 (L): Switchblade Legalization - Legalized the possession and carry of switchblade knives. 2017 (L): Constitutional Carry for Knives - Removed restrictions on carrying knives, including in a vehicle. 2018 (S): Extreme Risk Protection Orders - Implemented "red flag" law allowing temporary removal of firearms from individuals deemed a risk (Note: This may have faced legal challenges or implementation delays). 2021 (L): Second Amendment Sanctuary - Some counties declared themselves Second Amendment sanctuaries, though not at the state level. 2022 (L): Campus Carry Proposal - Significant discussions about allowing concealed carry on college campuses, though it may not have been enacted. 2023 (S): Universal Background Checks - Implemented background checks for private gun sales (Note: If not enacted, this was a significant proposal during this period). |
| Wyoming | 2007 (L): Castle Doctrine - Expanded self-defense rights in one's home and removed duty to retreat. 2010 (L): Firearms Freedom Act - Declared that firearms manufactured and retained in Wyoming are not subject to federal regulation. 2011 (L): Constitutional Carry - Implemented permitless carry, allowing legal gun owners to carry concealed firearms without a permit. 2013 (L): Hunting with Suppressors - Legalized the use of suppressors for hunting. 2015 (L): Campus Carry - Allowed concealed carry on college campuses for permit holders. 2017 (L): Stand Your Ground - Extended self-defense protections beyond the home to any place a person has a right to be. 2021 (L): Second Amendment Protection Act - Prohibited state and local officials from enforcing certain federal gun control measures. 2022 (L): Strengthened Preemption - Further reinforced state preemption of local firearm regulations, imposing penalties on local governments that enact gun restrictions. 2023 (L): Constitutional Carry Age Reduction - Lowered the age for permitless carry from 21 to 18 (Note: If not enacted, this was a significant proposal during this period). |

**Table S2**

| **State** | [2022 - Giffords Law Center Grade](https://giffords.org/lawcenter/resources/scorecard/) | [Everytown for Gun Safety Score](https://everytownresearch.org/rankings/) | [Brady Campaign Scores (2008)](https://www.gunpolicy.org/firearms/citation/quotes/7053) | [2021 - Gifford LC Grading](https://giffords.org/lawcenter/resources/scorecard/) | [2020 - Gifford LC Grading](https://giffords.org/lawcenter/resources/scorecard2020/#AK) | Grade |
| --- | --- | --- | --- | --- | --- | --- |
| Alabama | F | 12.50 | 15.00 | F | F | Most permissive |
| Alaska | F | 9.00 | 4.00 | F | F | Most permissive |
| Arizona | F | 8.50 | 6.00 | F | F | Most permissive |
| Arkansas | F | 9.50 | 6.00 | F | F | Most permissive |
| California | A | 89.50 | 79.00 | A | A | strict |
| Colorado | A- | 63.00 | 16.00 | B | C+ | Permissive |
| Connecticut | A | 82.50 | 54.00 | A- | A- | strict |
| Delaware | B+ | 61.50 | 22.00 | B | B | Permissive |
| Florida | D+ | 27.50 | 6.00 | C- | C- | Most permissive |
| Georgia | F | 5.00 | 7.00 | F | F | Most permissive |
| Hawaii | A- | 79.50 | 43.00 | A- | A- | strict |
| Idaho | F | 5.00 | 6.00 | F | F | Most permissive |
| Illinois | A- | 83.00 | 28.00 | A- | A- | strict |
| Indiana | D- | 16.50 | 8.00 | D- | D | Most permissive |
| Iowa | F | 15.50 | 16.00 | F | C | Most permissive |
| Kansas | F | 9.50 | 7.00 | F | F | Most permissive |
| Kentucky | F | 9.00 | 2.00 | F | F | Most permissive |
| Louisiana | F | 20.50 | 2.00 | F | F | Most permissive |
| Maine | D- | 20.50 | 12.00 | F | F | Most permissive |
| Maryland | A- | 75.00 | 53.00 | A- | A- | strict |
| Massachusetts | A- | 81.00 | 54.00 | A- | A- | strict |
| Michigan | B- | 35.00 | 22.00 | C+ | C | Permissive |
| Minnesota | B | 53.50 | 11.00 | C+ | C+ | Permissive |
| Mississippi | F | 3.00 | 5.00 | F | F | Most permissive |
| Missouri | F | 9.00 | 4.00 | F | F | Most permissive |
| Montana | F | 5.00 | 8.00 | F | F | Most permissive |
| Nebraska | C- | 25.00 | 10.00 | C- | C | Most permissive |
| Nevada | B- | 35.00 | 10.00 | C+ | C+ | Permissive |
| New Hampshire | D- | 9.00 | 11.00 | F | F | Most permissive |
| New Jersey | A | 79.00 | 63.00 | A | A | strict |
| New Mexico | C+ | 40.50 | 6.00 | C | C+ | Permissive |
| New York | A- | 83.50 | 51.00 | A- | A- | strict |
| North Carolina | C- | 25.00 | 20.00 | C- | D | Most permissive |
| North Dakota | F | 11.50 | 4.00 | F | F | Most permissive |
| Ohio | D- | 13.00 | 13.00 | D | D | Most permissive |
| Oklahoma | F | 7.50 | 2.00 | F | F | Most permissive |
| Oregon | A- | 68.00 | 18.00 | B- | C+ | Permissive |
| Pennsylvania | B | 40.00 | 26.00 | B- | C+ | Permissive |
| Rhode Island | B+ | 57.50 | 47.00 | B | B+ | strict |
| South Carolina | D- | 18.00 | 9.00 | F | F | Most permissive |
| South Dakota | F | 5.50 | 6.00 | F | F | Most permissive |
| Tennessee | F | 16.50 | 7.00 | F | D- | Most permissive |
| Texas | F | 13.50 | 9.00 | F | F | Most permissive |
| Utah | F | 12.00 | 4.00 | F | D | Most permissive |
| Vermont | B- | 39.50 | 9.00 | C- | C- | Permissive |
| Virginia | B+ | 49.00 | 18.00 | B | B | Permissive |
| Washington | A- | 69.00 | 18.00 | B | B+ | Permissive |
| West Virginia | F | 18.50 | 4.00 | F | F | Most permissive |
| Wisconsin | C | 28.00 | 12.00 | C- | C- | Most permissive |
| Wyoming | F | 6.50 | 9.00 | F | F | Most permissive |

**Figure S1. Pediatric firearm mortality (all intents) by legal grouping with yearly data shown.**

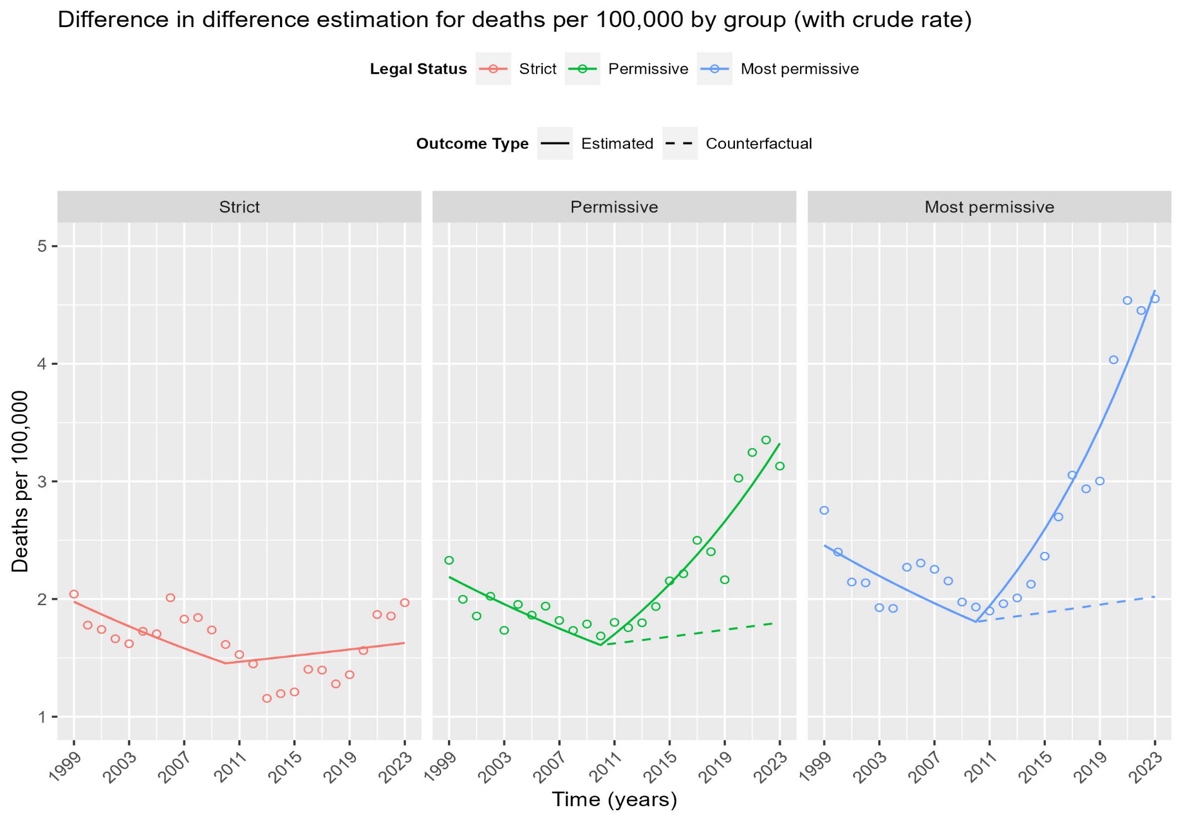

| **State groups** | **Total # of Excess deaths compared to strict group (post policy)** | **Total observed # of deaths from the model (post policy)** | **Total counterfactual # of deaths from the model (post policy)** | **Overall excess mortality rate (per 100,000, post policy)** | **Interaction p-value** |
| --- | --- | --- | --- | --- | --- |
| Permissive | 1,236 | 4,180 | 2,944 | 0.72 | <0.001 |
| Most permissive | 5,893 | 15,378 | 9,485 | 1.19 | <0.001 |

**Figure S2. Pediatric neoplasms mortality by firearm legal grouping sensitivity analysis.**

**Panel A**

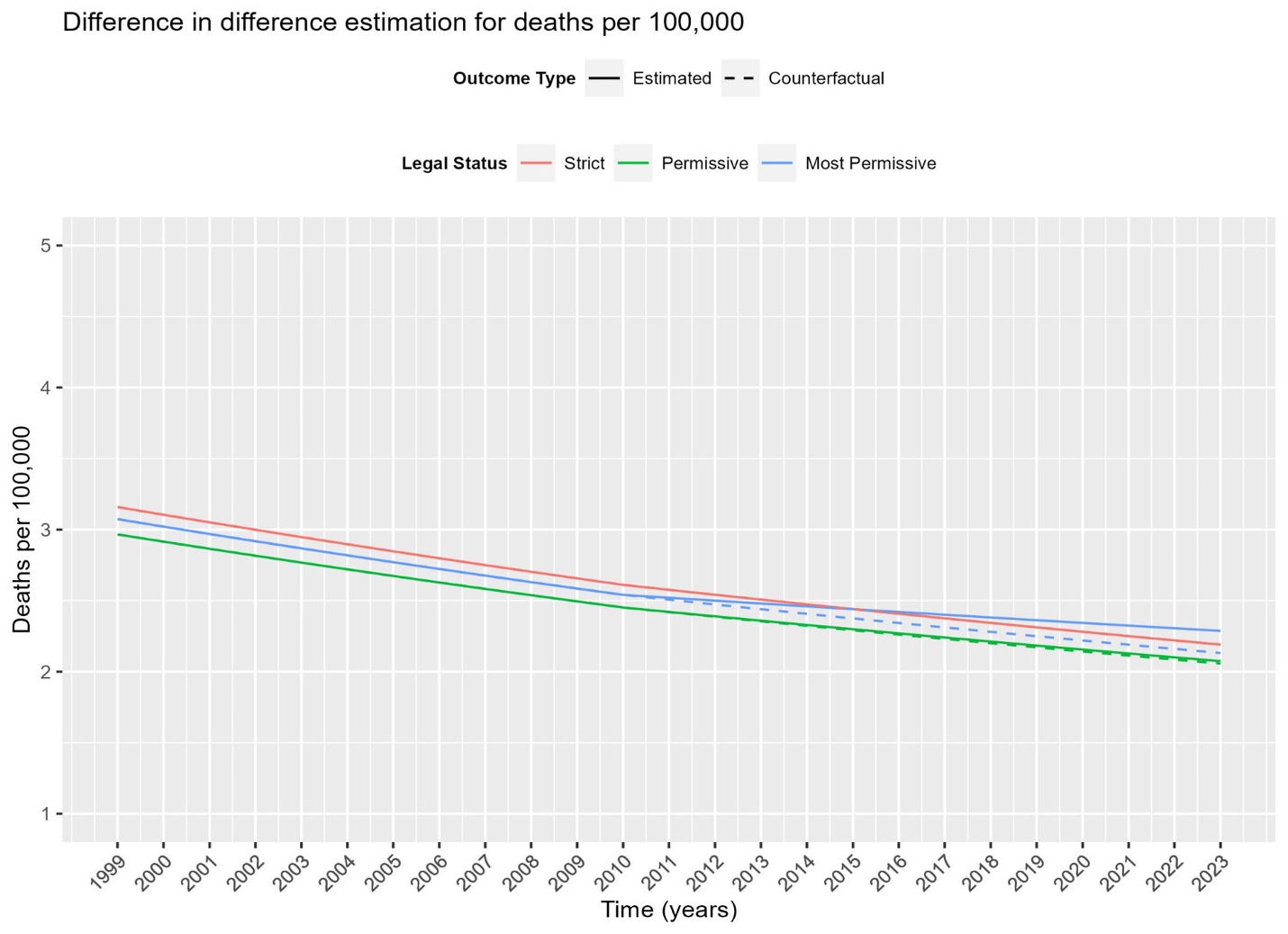

| **State groups** | **Total # of Excess deaths compared to strict group (post policy)** | **Total observed # of deaths from the model (post policy)** | **Total counterfactual # of deaths from the model (post policy)** | **Overall excess mortality rate (per 100,000, post policy)** | **Interaction p-value** |
| --- | --- | --- | --- | --- | --- |
| Permissive | 17 | 3,864 | 3,848 | 0.01 | 0.844 |
| Most Permissive | 434 | 11,866 | 11,432 | 0.09 | 0.027 |

**Panel** **B. Pediatric neoplasms mortality by firearm legal grouping with yearly data.**

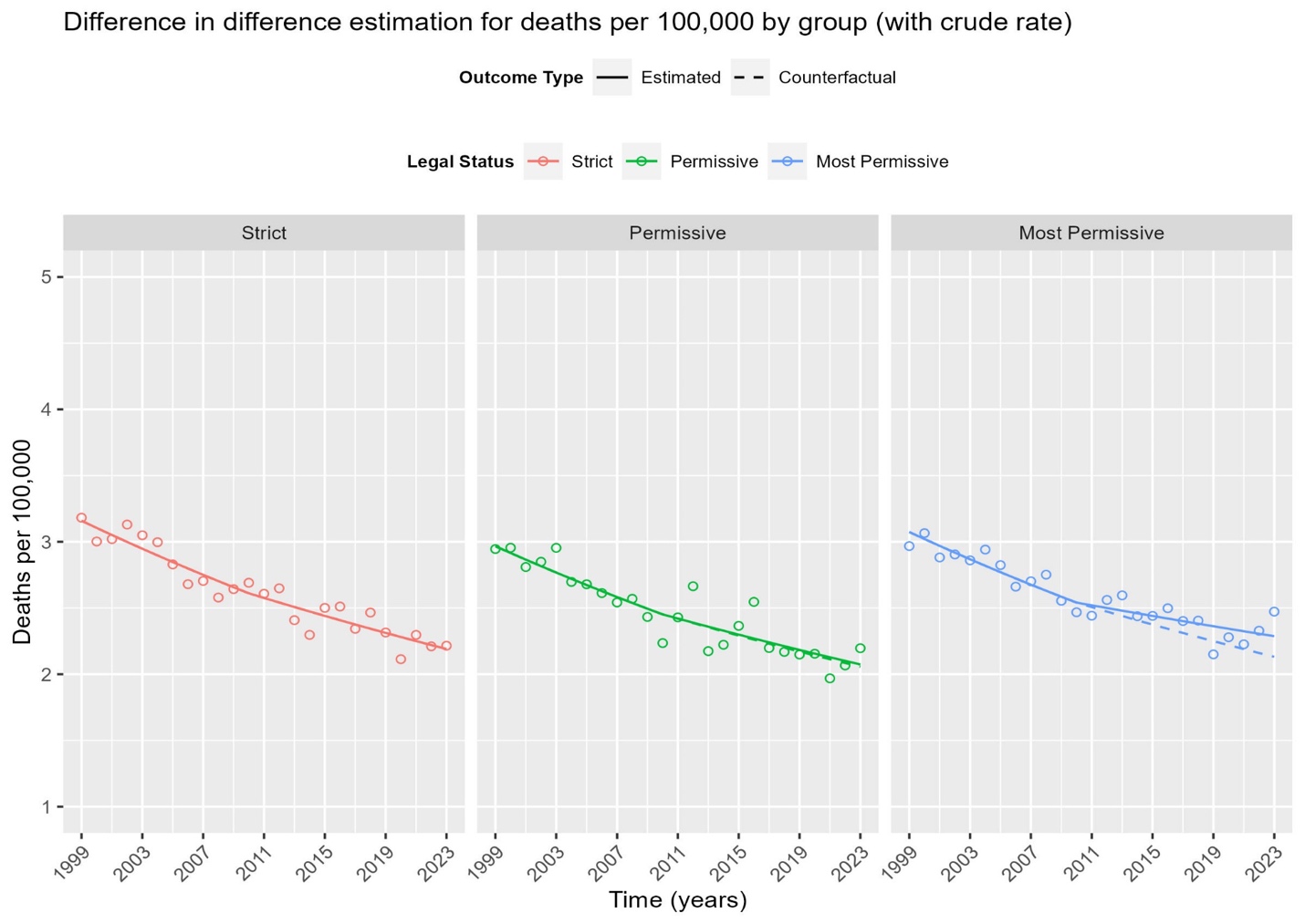

**Figure S3. Pediatric motor vehicle mortality by firearm legal grouping sensitivity analysis.**

**Panel A**
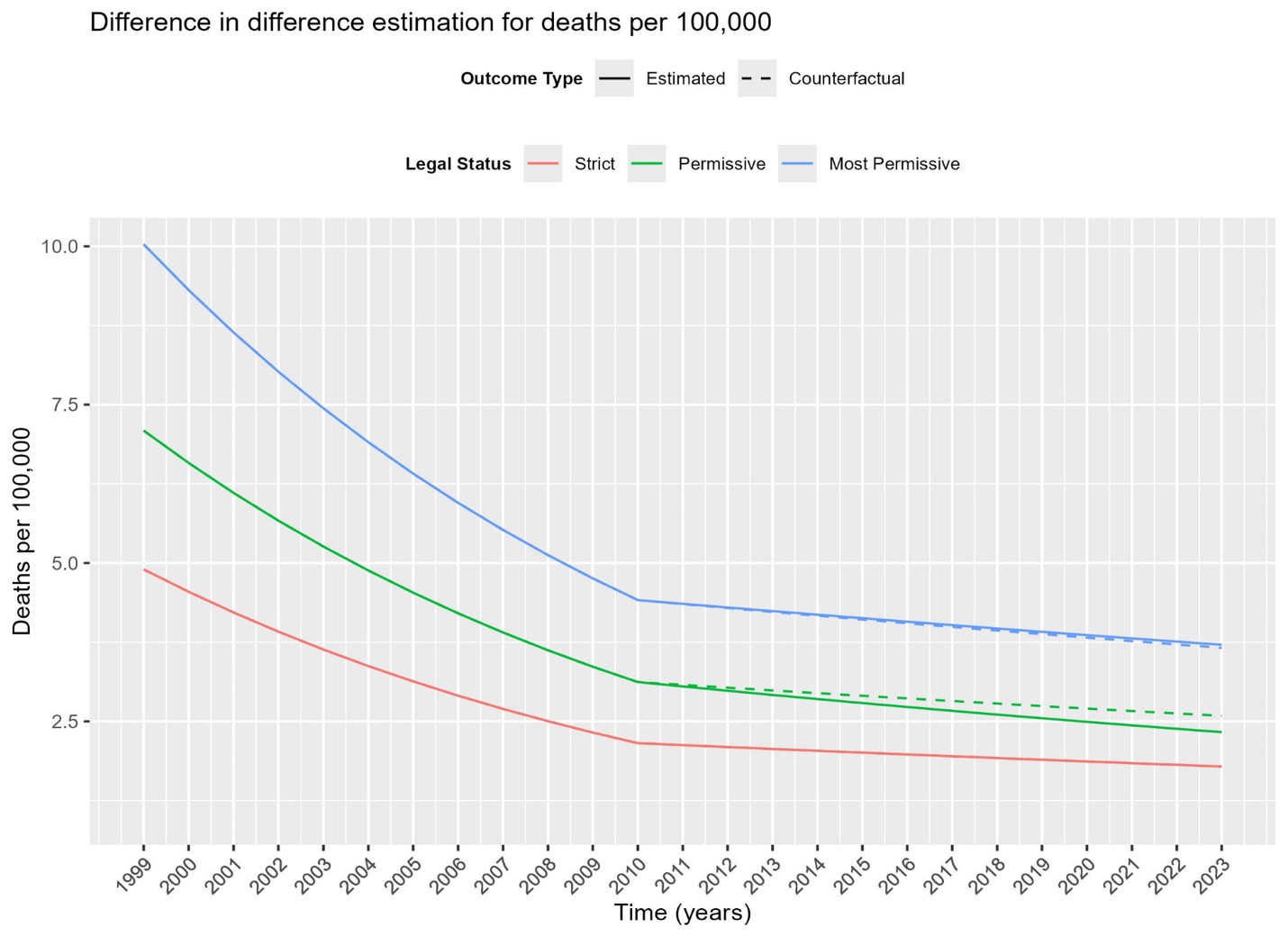

| **State groups** | **Total # of Excess deaths compared to strict group (post policy)** | **Total observed # of deaths from the model (post policy)** | **Total counterfactual # of deaths from the model (post policy)** | **Overall excess mortality rate (per 100,000, post policy)** | **Interaction p-value** |
| --- | --- | --- | --- | --- | --- |
| Permissive | -257 | 4,610 | 4,867 | -0.15 | 0.009 |
| Most Permissive | 137 | 19,880 | 19,743 | 0.03 | 0.664 |

**Panel B. Pediatric motor vehicle mortality by firearm legal grouping sensitivity analysis,**

**yearly data.**

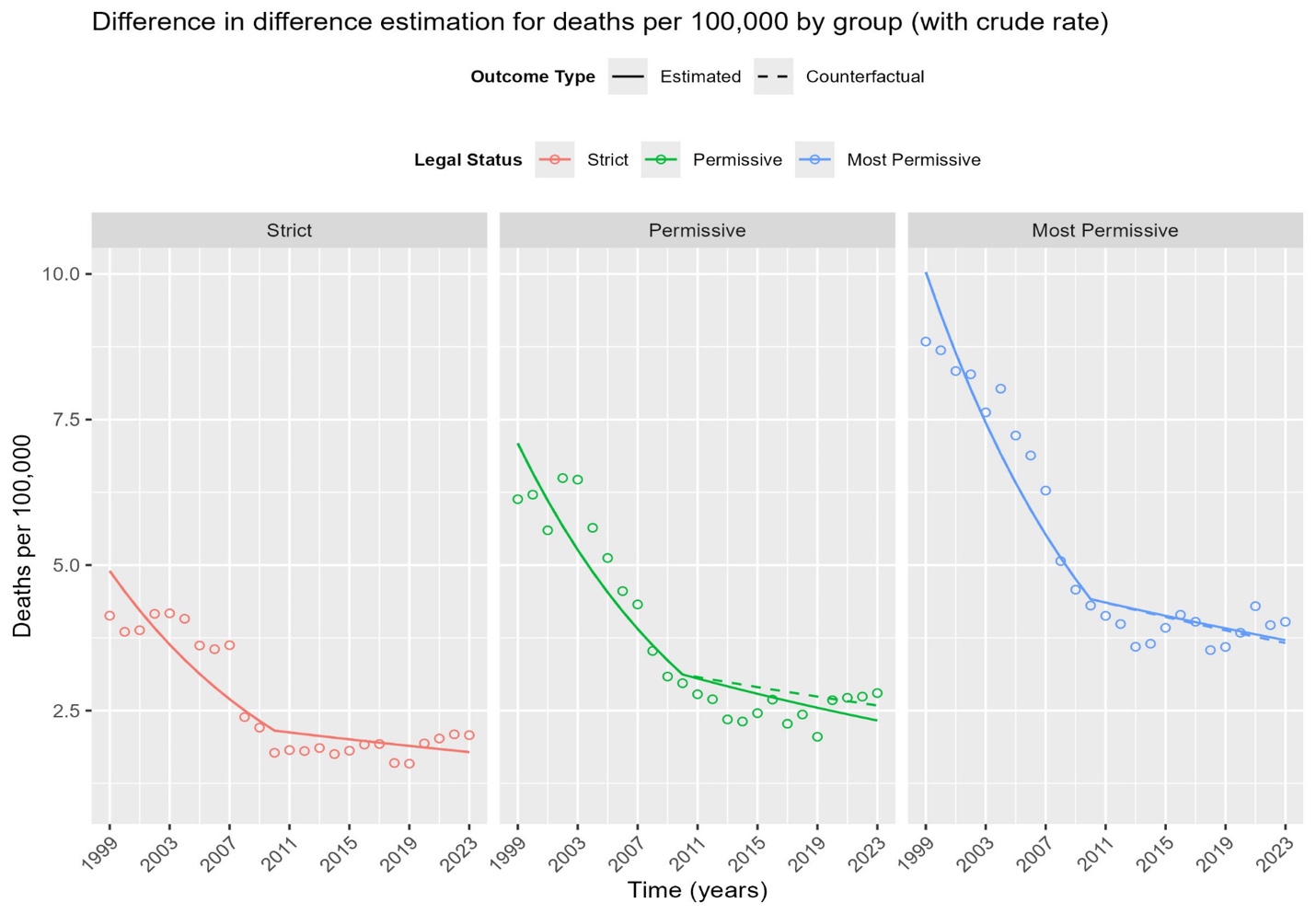

**Figure S4. Pediatric homicide firearm deaths by firearm legal grouping with yearly data.**

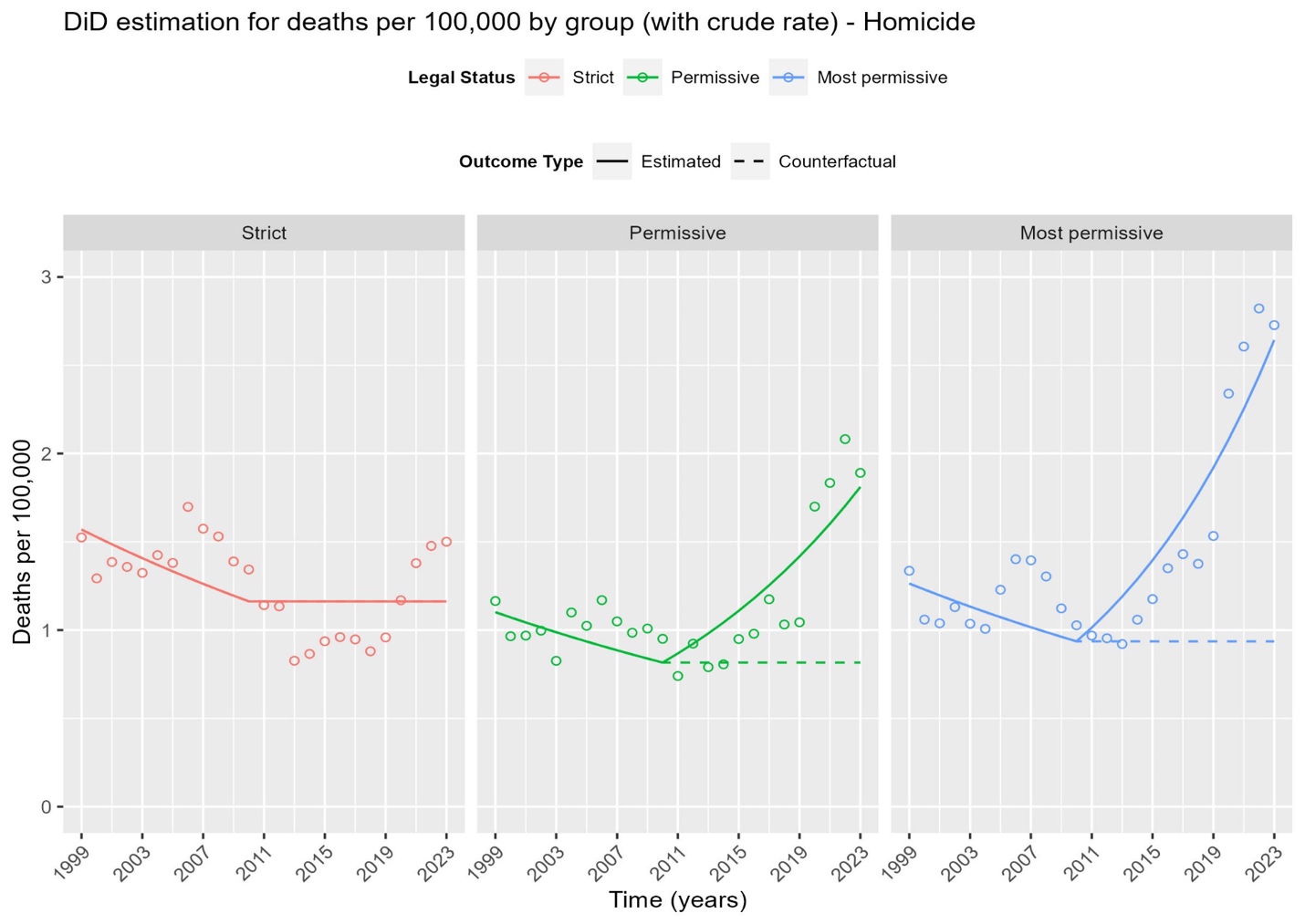

| **State groups** | **Total # of Excess deaths compared to strict group (post policy)** | **Total observed # of deaths from the model (post policy)** | **Total counterfactual # of deaths from the model (post policy)** | **Overall excess mortality rate (per 100,000, post policy)** | **Interaction p-value** |
| --- | --- | --- | --- | --- | --- |
| Permissive | 808 | 2,213 | 1,405 | 0.47 | <0.001 |
| Most permissive | 3,840 | 8,465 | 4,625 | 0.78 | <0.001 |

**Figure S5. Pediatric suicide firearm deaths by firearm legal grouping with yearly data.**

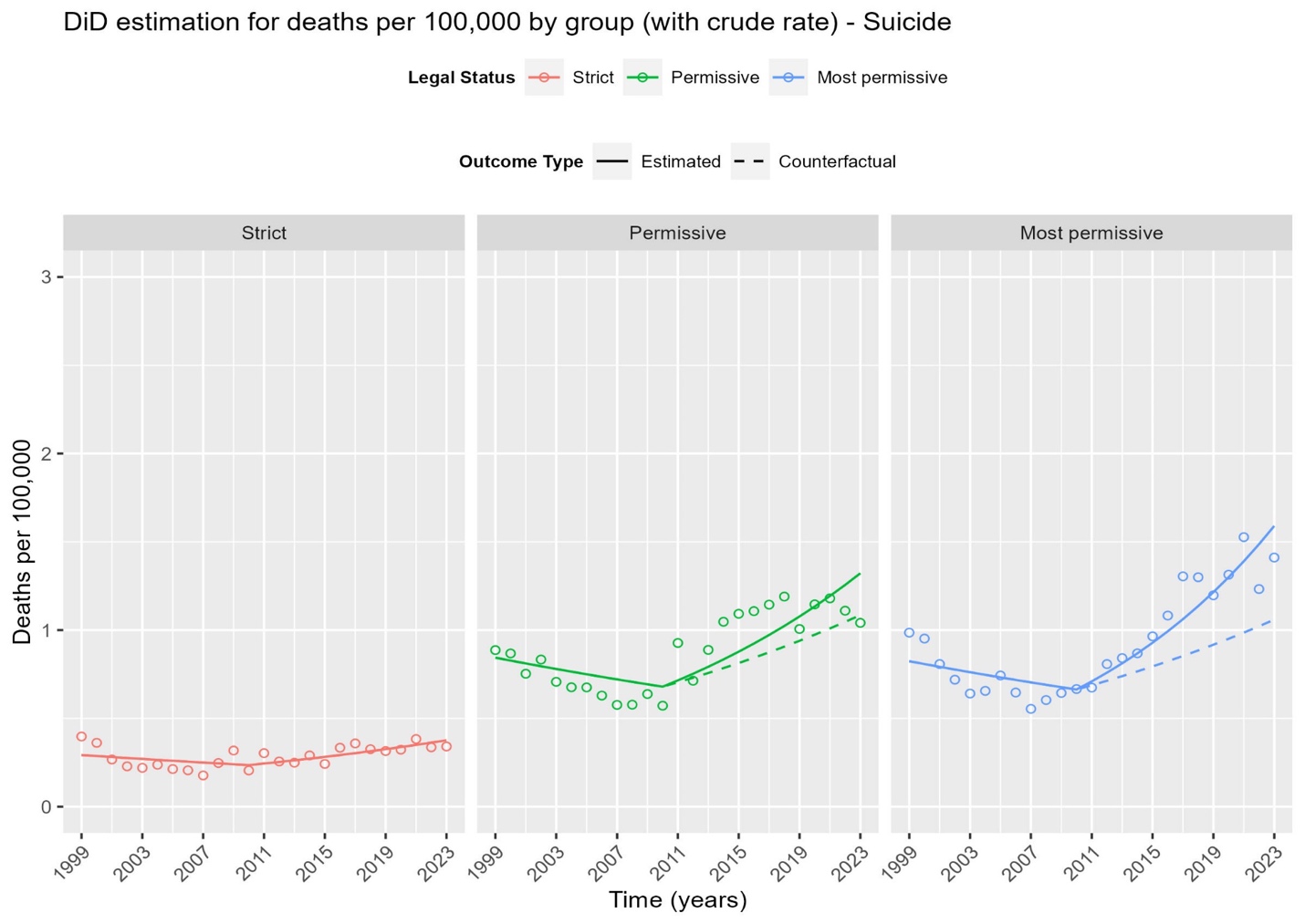

| **State groups** | **Total # of Excess deaths compared to strict group (post policy)** | **Total observed # of deaths from the model (post policy)** | **Total counterfactual # of deaths from the model (post policy)** | **Overall excess mortality rate (per 100,000, post policy)** | **Interaction p-value** |
| --- | --- | --- | --- | --- | --- |
| Permissive | 186 | 1,703 | 1,518 | 0.11 | 0.023 |
| Most permissive | 1,164 | 5,422 | 4,258 | 0.24 | <0.001 |

**Figure S6. Pediatric firearm mortality by 2013 urbanicity, most permissive states, yearly data.**

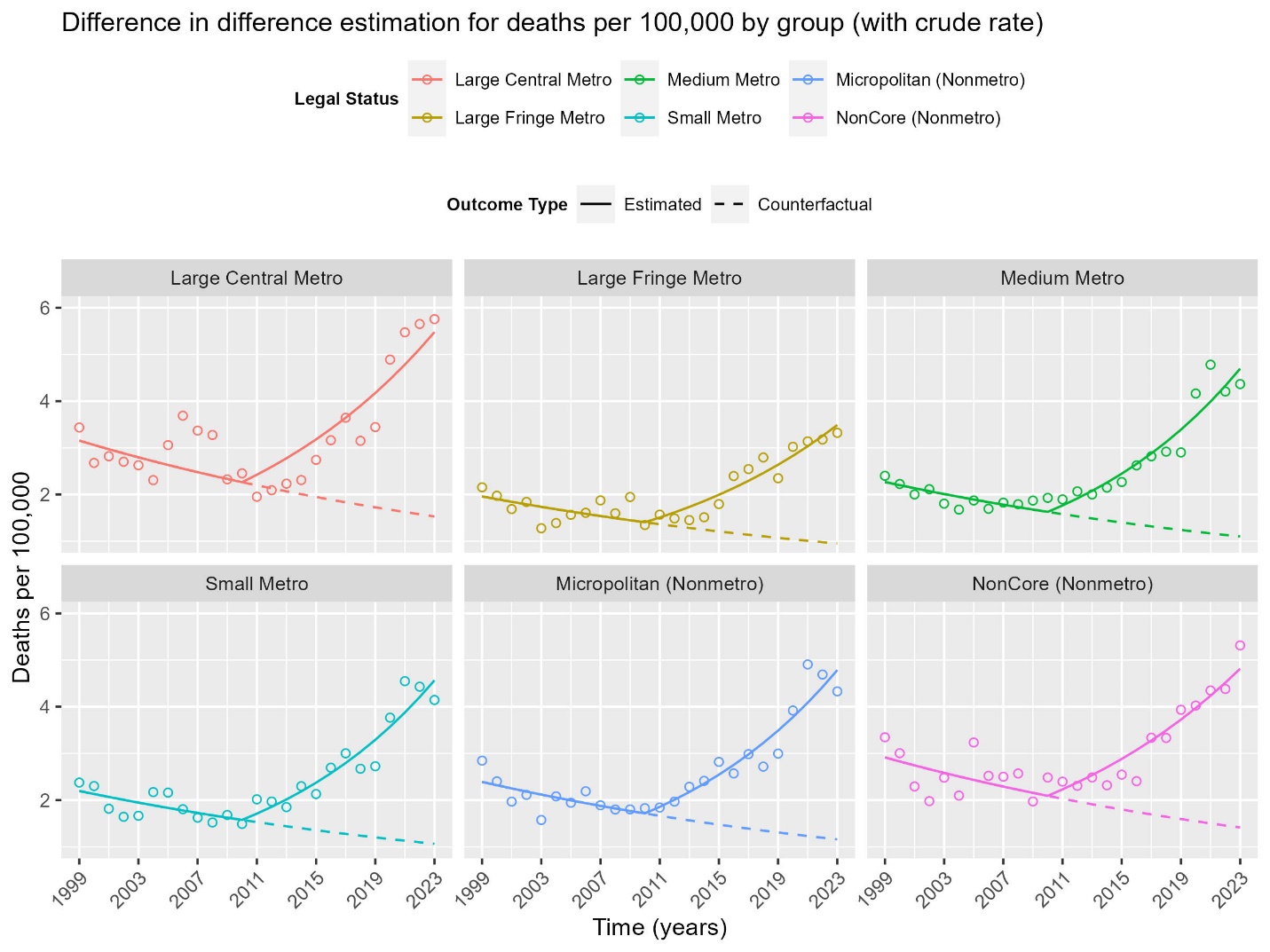

| **Urbanicity groups (2013)** | **Total # of Excess deaths compared to strict group (post policy)** | **Total observed # of deaths from the model (post policy)** | **Total counterfactual # of deaths from the model (post policy)** | **Overall excess mortality rate (per 100,000, post policy)** | **Interaction p-value** |
| --- | --- | --- | --- | --- | --- |
| Large Central Metro | 2,381 | 4,653 | 2,272 | 1.93 | <0.001 |
| Large Fringe Metro | 1,212 | 2,326 | 1,114 | 1.24 |  |
| Medium Metro | 2,060 | 3,672 | 1,612 | 1.69 |  |
| Small Metro | 902 | 1,607 | 705 | 1.64 |  |
| Micropolitan (Nonmetro) | 933 | 1,702 | 769 | 1.70 |  |
| NonCore (Nonmetro) | 682 | 1,389 | 708 | 1.65 |  |

**Figure S7. Pediatric firearm deaths by observed race/ethnicity, most permissive states, yearly data.**

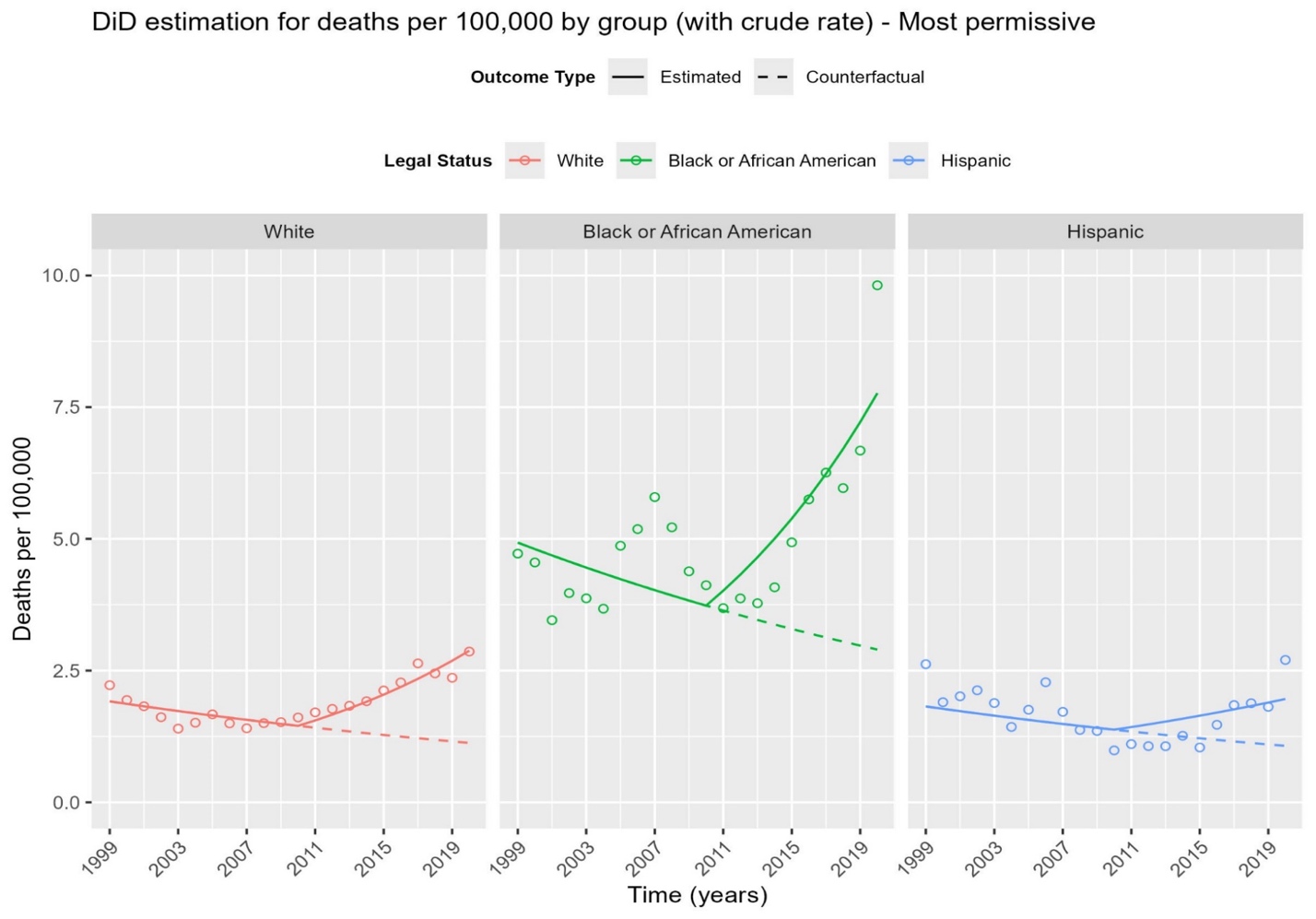

| **Race/ethnicity groups** | **Total # of Excess deaths compared to strict group (post policy)** | **Total observed # of deaths from the model (post policy)** | **Total counterfactual # of deaths from the model (post policy)** | **Overall excess mortality rate (per 100,000, post policy)** | **Interaction p-value** | **p-value for mortality slope change** |
| --- | --- | --- | --- | --- | --- | --- |
| White | 1,898 | 4,624 | 2,726 | 0.88 | <0.001 | <0.001 |
| Black or African American | 1,634 | 3,797 | 2,163 | 2.46 |  | <0.001 |
| Hispanic | 399 | 1,374 | 975 | 0.49 |  | <0.001 |
